# A One-Shot Lossless Algorithm for Cross-Cohort Learning in Mixed-Outcomes Analysis

**DOI:** 10.1101/2024.01.09.24301073

**Authors:** Ruowang Li, Luke Benz, Rui Duan, Joshua C. Denny, Hakon Hakonarson, Jonathan D. Mosley, Jordan W. Smoller, Wei-Qi Wei, Thomas Lumley, Marylyn D. Ritchie, Jason H. Moore, Yong Chen

## Abstract

In cross-cohort studies, integrating diverse datasets, such as electronic health records (EHRs), is both essential and challenging due to cohort-specific variations, distributed data storage, and data privacy concerns. Traditional methods often require data pooling or complex data harmonization, which can reduce efficiency and limit the scope of cross-cohort learning. We introduce mixWAS, a one-shot, lossless algorithm that efficiently integrates distributed EHR datasets via summary statistics. Unlike existing approaches, mixWAS preserves cohort-specific covariate associations and supports simultaneous mixed-outcome analyses. Simulations demonstrate that mixWAS outperforms conventional methods in accuracy and efficiency across various scenarios. Applied to EHR data from seven cohorts in the US, mixWAS identified 4,530 significant cross-cohort genetic associations among traits such as blood lipids, BMI, and circulatory diseases. Validation with an independent UK EHR dataset confirmed 97.7% of these associations, underscoring the algorithm’s robustness. By enabling lossless cross-cohort integration, mixWAS improves the precision of multi-outcome analyses and expands the potential for actionable insights in healthcare research.

**The bigger picture:** Cross-cohort integration of electronic health record (EHR) datasets is critical for advancing genomic discovery but remains hindered by privacy concerns, cohort heterogeneity, and computational limitations. Traditional meta-analysis and federated methods either lose power or cannot fully model multiple mixed-outcome traits across distributed datasets. To address this, we developed mixWAS, a one-shot, lossless algorithm for integrating summary statistics across cohorts without sharing individual-level data. mixWAS simultaneously models binary and continuous outcomes, accounts for site-specific covariate heterogeneity, and requires only a single communication step between sites. Through extensive simulations and real data analyses, mixWAS consistently outperformed traditional Phenome-Wide Association Studies (PheWAS) and other multi-trait approaches in detecting multi-phenotype associations (MPAs). eyond genetic applications, mixWAS offers a general framework for distributed analysis of mixed-outcome data, with broad potential across biomedicine, public health, and other fields requiring privacy-preserving data integration.

**Highlights:**

- mixWAS enables lossless, one-shot cross-cohort integration of summary statistics
- Simultaneously models binary and continuous outcomes across distributed datasets
- Outperforms PheWAS in detecting multi-phenotype associations (MPA)
- Offers a general framework for distributed analysis of mixed-outcome data,

## Introduction

Cross-cohort studies increasingly require robust methods to integrate heterogeneous datasets, especially in contexts where multiple outcomes—such as phenotypic traits, clinical measures, and other health indicators—are examined concurrently. The increasing availability of biobank-linked electronic health record data (EHR), such as the UK Biobank (UKBB) and Electronic Medical Records and Genomics Network (eMERGE), provides a rich source for uncovering complex multi-outcome associations. For example, multi-phenotype associations (MPA), where a single genetic variant is linked to multiple traits, reveal potential shared genes and pathways across diseases. Since patients’ genetic data can be mapped to a wide range of clinically relevant phenotypes, these datasets enhance opportunities for comprehensive multi-outcome analysis^1–9^.

Genome-wide association studies (GWAS) have systematically identified numerous genetic associations for various diseases and traits^10–12^,yet challenges persist in uncovering shared genetic and non-genetic architectures across multiple outcomes^13–15^. Many observed multi-outcome associations ^16–18^, such as MPA, reveal complex genetic relationships across traits and may provide insights into disease pathogenesis^17,19–22^. Moreover, identifying MPA holds significant clinical values across various domains, including the development of targeted therapies against genes or pathways involved in the development of multiple diseases^23^, repurposing existing drugs targeting shared druggable genes across diseases^24^, and improving disease risk prediction and screening by incorporating information about share genetic factors across diseases^25,26^. Thus, identifying MPA is a crucial next step towards improved understanding of the genetic architectures of complex diseases.

The integration of multiple EHRs for cross-cohort analysis improves the power to detect MPA and improves the reproducibility of findings. However, current computational methodologies fall short in fully leveraging distributed EHR data for MPA discovery. Due to the distributed nature of EHR systems, individual-level data often cannot be pooled across sites for integrated analysis. As a result, summary-level integration approaches such as meta-analysis and federated learning algorithms are commonly employed for cross-cohort studies. Phenome-Wide Association Studies (PheWAS)—which conduct separate GWAS for each phenotype followed by combination of results—are widely used for MPA detection. Meta-analysis has become the standard for integrating PheWAS findings across EHR datasets to boost power^22,27–30^. However, PheWAS suffers from a loss of power due to stringent multiple testing corrections that arise from the large number of variant-phenotype combinations ^31,32^. Furthermore, meta-analyzed PheWAS results often differ from those obtained via pooled analysis and have been shown to be less powerful in detecting MPAs across EHR cohorts^1,30,33^. The sPLINK framework offers a federated alternative to meta-analysis, accounting for cross-study heterogeneity in GWAS without pooling individual-level data^34^. Yet, sPLINK is limited to single-phenotype analyses and does not resolve the multiple testing burden inherent in PheWAS. Similarly, FedGMMAT extends generalized linear mixed models to a federated setting for binary traits but remains constrained to individual phenotype analysis^35^. SF-GWAS, based on REGENIE’s stacked ridge regression approach, is another distributed method for GWAS of individual traits^36^. Alternative approaches such as ASSET^37^, GPN^38^, MTAG^39^, MultiPhen^40^, and subSETs^41^ offer more efficient multi-trait testing strategies, but they are typically limited to either binary or continuous phenotypes and often only applicable to individual datasets^22,42^. In practice, EHR-derived phenotypes frequently consist of a mixture of continuous and binary outcomes. Therefore, there is a critical need for a federated, lossless method that can jointly model mixed-outcome phenotypes across distributed EHR datasets to detect MPAs more effectively.

To address these issues, we introduce *mixWAS*, a one-shot lossless algorithm designed for versatile cross-cohort integration, particularly suited to identifying associations across diverse, mixed-outcome datasets. We implemented four key features in mixWAS to fulfill these goals. First, mixWAS can identify genetic associations among mixed-type phenotypes, including binary (e.g., case-control) and continuous (e.g., lab measurements). Second, mixWAS can efficiently and losslessly integrate multiple EHRs to increase the overall sample size. Third, mixWAS is designed to handle phenotype and confounding covariate heterogeneity, such as site-specific covariate associations, that may exist among different EHRs. For example, different age-onset for different diseases or block-wise missing data in some EHRs. Finally, mixWAS only requires data summary statistics from different EHRs, minimizing data transferring and communication costs.

Using simulations, we first demonstrated that the mixWAS algorithm has better power than the commonly used PheWAS approach. Subsequently, the proposed method was utilized to detect MPA among cardiovascular related mixed-type phenotypes using patients from seven EHR data from eMERGE. We then validated our findings in the independent UKBB data. In total, mixWAS identified 4,530 associations in the integrated analysis using all eMERGE EHRs of which 4,428 SNPs (97.7%) were validated by the independent UKBB data. In summary, the proposed mixWAS algorithm can efficiently integrate EHR data from multiple sources to detect MPA among mixed phenotypes, paving the way for discovering new insights into disease mechanisms and potential therapeutic targets. Beyond genetic data, the algorithm also has broad applicability to any distributed real-world datasets with mixed outcome types.

## Results

### Overview of the mixWAS algorithm

mixWAS is designed to identify genetic variants associated with multiple diseases or traits using distributed datasets. It is specifically tailored to handle mixed phenotypes, including both binary and continuous outcomes. A central challenge in jointly analyzing such mixed outcomes is the specification of their joint distribution, which is often unknown or complex. Consequently, most existing MPA approaches either analyze one outcome at a time or jointly analyze outcomes of the same type. mixWAS overcomes this limitation by employing a composite likelihood framework combined with a robust sandwich variance estimator. This approach eliminates the need to specify joint distributions, enabling effective integration of evidence for MPAs across a wide range of phenotype types. In addition, mixWAS accounts for site-specific covariate effects, thereby addressing distributional shifts in exposure variables commonly observed across EHR datasets. Such shifts are a significant challenge in real-world data analysis and are expected when working with heterogeneous EHR sources^43^. Unlike meta-analysis, which often yields results that differ from pooled individual-level analyses and may lose power due to data summarization, mixWAS enables lossless integration across datasets. Its design allows for the effective and efficient use of distributed data while achieving results identical to pooled individual-level analyses. The algorithm follows a three-step process: First, intermediate summary statistics of genetic associations are calculated within each distributed dataset. These summary statistics are then transmitted to a centralized server and analyzed by an analyst, who computes the components (score and variance) of the test statistic. Finally, the test statistics are derived, yielding p-values for subsequent inference (Figure 1).

**Figure 1:**
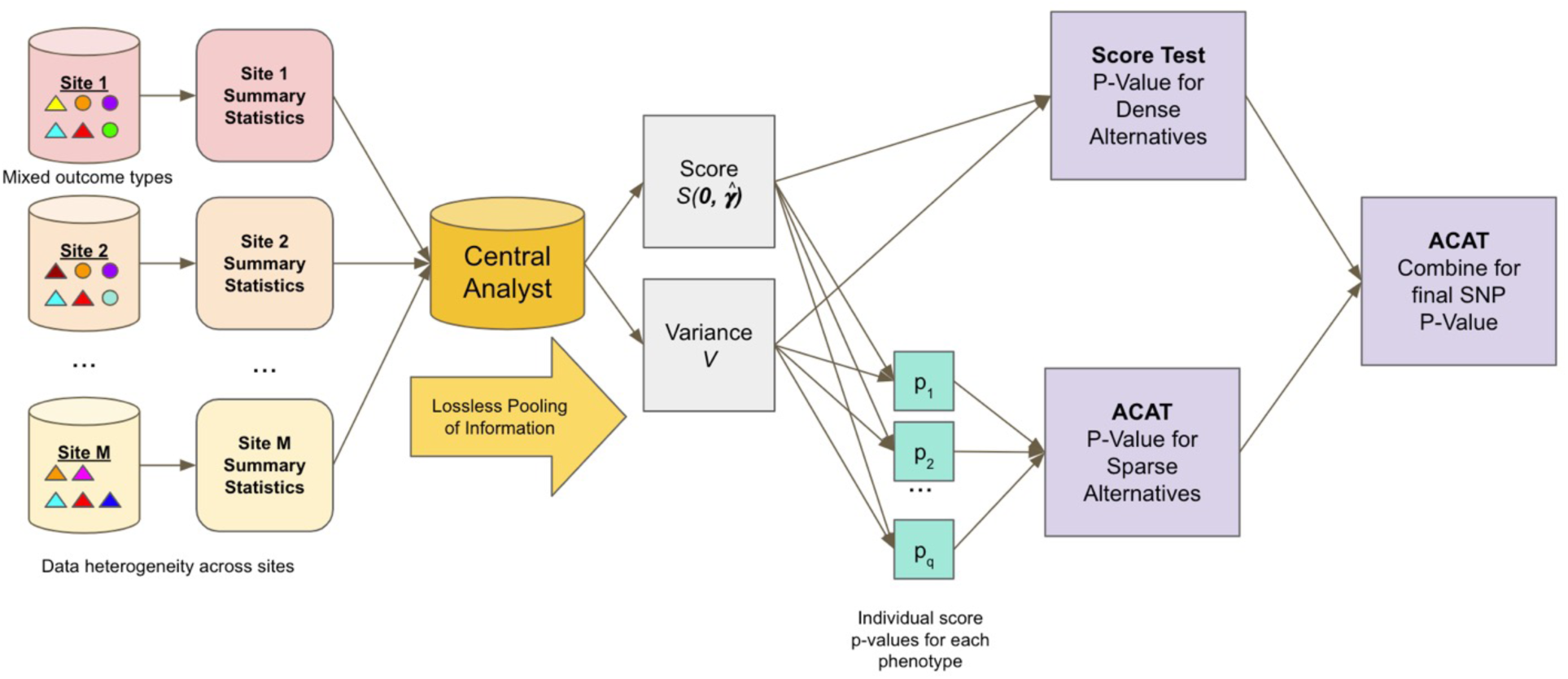
Outline of the mixWAS algorithm. Each site transmits summary level statistics of heterogeneous, mixed-typed phenotypes, specifically the score vector *S_m_*(**0**, ***γ***_m_) and variance matrix *V_m_*, to a central analyst. The central analyst pools the site-specific score and variance contributions in a lossless manner for use in a score test, which is powerful against dense phenotypes, and a second test, robust to sparse phenotypes, which combines the *p*-values of individual score tests for each of *q* phenotypes using the ACAT method^44^. Finally, these two *p*- values, one optimized against dense phenotypes and the other optimized against sparse phenotypes, are combined again using the ACAT method.

### mixWAS is more powerful than PheWAS and MultiPhen in detecting MPA across mixed- type phenotypes

PheWAS is the most commonly used method to detect MPA. Unlike other methods for detecting MPA, PheWAS can be readily extended to handle mixed phenotypes in multiple datasets through meta-analysis. Thus, mixWAS can directly be compared to PheWAS to evaluate its power to detect MPA. MultiPhen^40^ is another common method for detecting MPA, by using phenotypes as covariates to model the SNP as an outcome. As implemented, MultiPhen is less directly comparable to mixWAS than traditional PheWAS methods because beta estimates for each phenotype don’t have associated standard errors, and thus can’t be combined across sites via standard meta-analysis techniques. To evaluate MultiPhen in our setting, we devised a simple federated version of MultiPhen, described in more detail below.

In order to compare mixWAS to various PheWAS methods, as well as multiPhen we conducted various simulation studies across many diverse settings. Each simulation setting consisted of five independent datasets representing distributed sites. Within each dataset, we simulated MPA associations and confounding covariates, including principal components, age, and sex. Covariate effects for each phenotype were intentionally varied across datasets to reflect heterogeneity across sites. The true MPA associations were devised to two distinct scenarios in simulation studies: 1) Same direction: all phenotypes (binary and continuous) were positively or negatively associated with the genotype in the same direction (Figure 2A), and 2) Opposite direction: half of the phenotypes had positive associations with the genotype and half had negative associations (Figure 2B). We also incorporated additional factors such as signal sparsity, phenotype correlations, missing data, and different ratios of mixed phenotypes in our simulations.

**Figure 2:**
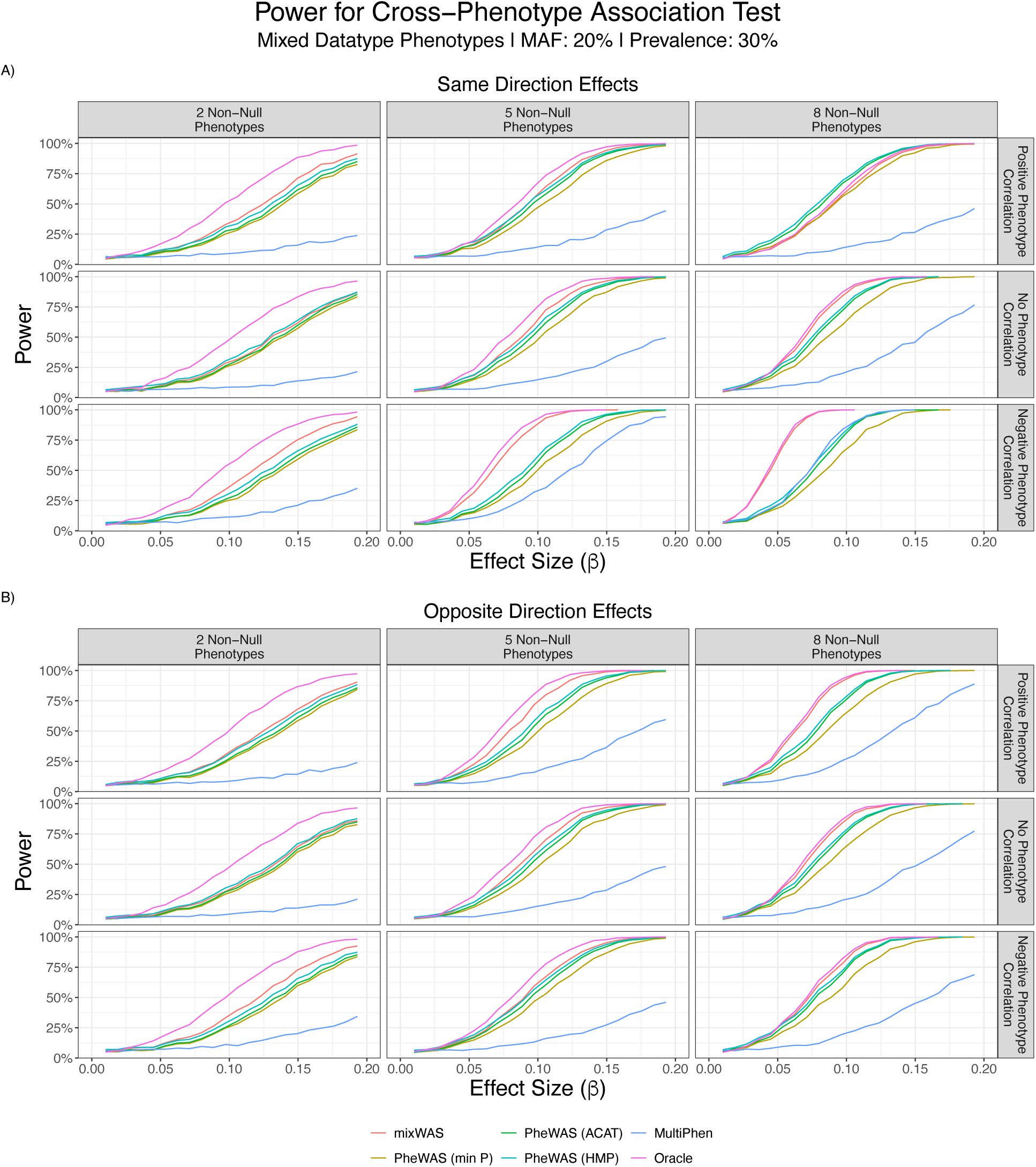
Empirical power curves comparing various cross-phenotype association tests for simulated mixed-type phenotypes. **A)** The simulated SNP is positively associated with all phenotypes, while the MPA sparsity (e.g. number of phenotypes with significant association) and correlation between phenotypes vary. **B)** The simulated SNP is positively associated with all binary phenotypes and is positively associated with half of the continuous phenotypes and negatively associated with the other half. MPA sparsity and correlation between phenotypes vary.

The simulated data was analyzed using mixWAS, PheWAS, and multiPhen, as described briefly below. mixWAS generated intermediate summary-level statistics in each dataset and performed integration using these statistics to calculate the association p-values. In contrast, separate PheWAS analyses were conducted in each dataset under PheWAS, where the beta coefficients were combined using meta-analysis. A final PheWAS p-value was obtained using three distinct methods: 1) taking the minimal p-value across phenotypes and applying Bonferroni correction; 2) harmonic mean p-value (HMP)^45^; 3) ACAT. The federated version of MultiPhen conducted a single analysis at each site and obtained a final p-value by taking the minimum p-value of the joint model from each site and applying Bonferroni correction for the number of sites. Finally, we ascertained the highest possible performance of any score-based test using an oracle model, by applying the score test for dense alternatives in the true subset of phenotypes associated with the SNP.

Across all simulation settings, mixWAS consistently outperformed PheWAS and MultiPhen (Figure 2). The gains in power by using mixWAS over other methods were greatest when the direction of SNP effects went against the direction of the residual correlation between phenotypes. For example, mixWAS outperformed PheWAS the most when SNP effects were positive and residual correlation was negative reflective of a setting where positive correlation genetic correlation exists among traits in the presence of negative environmental or other correlation (Figure 2A). mixWAS also outperformed PheWAS methods when SNP effects had opposite signs with positive residual phenotype correlation (Figure 2B). Such power gains result from the fact that unlike PheWAS methods, mixWAS accounts for correlation between phenotypes. The heterogeneity in MPA effects and the residual correlations are frequently encountered in practical settings, underscoring the practical usefulness of mixWAS.

PheWAS methods combining p-values via ACAT or HMP obtained higher power than any possible score-based test in the fully dense setting with positive phenotype correlation, but the power of all PheWAS methods were dramatically influenced by the direction of correlation. By contrast, the power of mixWAS fluctuated much less when changing correlation direction. MultiPhen performed significantly worse than either mixWAS or PheWAS across settings. Missing phenotypes present a significant challenge for MultiPhen, which uses them as covariates and thus drops any subject with a single missing phenotype, unlike mixWAS, which accounts for missingness in the score, or PheWAS, which drops that subject from analysis of the corresponding phenotype but retains that subject for analysis of additional non-missing phenotypes. Type 1 errors were controlled at the nominal level for all methods.

Additional simulations on binary-only phenotypes (Supplemental Figure S4), designs using shared healthy controls (Supplemental Figure S5), missing at random outcomes (Supplemental Figure S7), and heterogenous covariate distributions across sites (Supplemental Figure S8) further demonstrated the superior power of mixWAS compared to PheWAS and MultPhen.

### Detecting MPA across blood lipids levels, BMI, and diseases of circulatory system

Previous research has identified MPA between BMI and coronary heart disease ^46,47^, blood lipids levels^48^, and low-density lipoprotein, triglycerides, and cardiovascular diseases^49^. These studies have highlighted the existence of potential shared underlying genetic architecture among these traits/diseases. However, comprehensive investigations of MPA across all traits and diseases have not been carried out. Leveraging multiple EHR datasets and the improved efficiency of our proposed method, we utilized mixWAS to detect MPA among blood lipid levels (high-density lipoprotein (HDL), low-density lipoprotein (LDL), serum total cholesterol, and triglycerides), body mass index (BMI), and circulatory diseases (unspecified essential hypertension, type 2 diabetes (T2D), unspecified hyperlipidemia, benign essential hypertension, atrial fibrillation, congestive heart failure, and coronary atherosclerosis) using eMERGE data from 7 sites. The characteristics of the datasets are presented in Supplemental Figure S1, illustrating heterogeneities among the EHRs, including variations in the number of patients and patterns of missing data.

To identify MPA, we conducted three separate analyses: eMERGE single-site analysis, eMERGE integrated analysis via mixWAS, and external validation using the UKBB dataset. In all analyses, patients’ sex, ten principal components accounting for population stratification, and trait-associated ages were adjusted in the model. Notably, each trait was measured at a different associated age, leading to distinctive adjustments for each trait. In the first analysis, mixWAS was individually applied to each eMERGE dataset to detect MPA. This allowed the assessment of the power of detecting MPA when each dataset was used independently. The Manhattan plots showed that datasets with smaller sizes (Marshfield, Northwestern, Geisinger, and Kaiser Permanente) exhibited lower power in detecting MPA compared to larger datasets (Mass General Brigham, Vanderbilt, and Mayo). Next, an integrated analysis was performed by mixWAS utilizing all eMERGE datasets. This integrated analysis significantly increased the total sample size and identified a higher number of significant MPA compared to any individual dataset. The Bonferroni-corrected significant associations identified in the eMERGE integrated analysis were validated using the independent UK Biobank data. Out of 4,530 eMERGE associations, 4,428 (97.7%) were successfully replicated in the UKBB dataset, providing further validation for the identified associations (Figure 3 and Supplemental Table S1). In contrast, meta-analyzed PheWAS applied across all eMERGE datasets has yielded only 98 significant associations (Supplemental Table S3).

**Figure 3.**
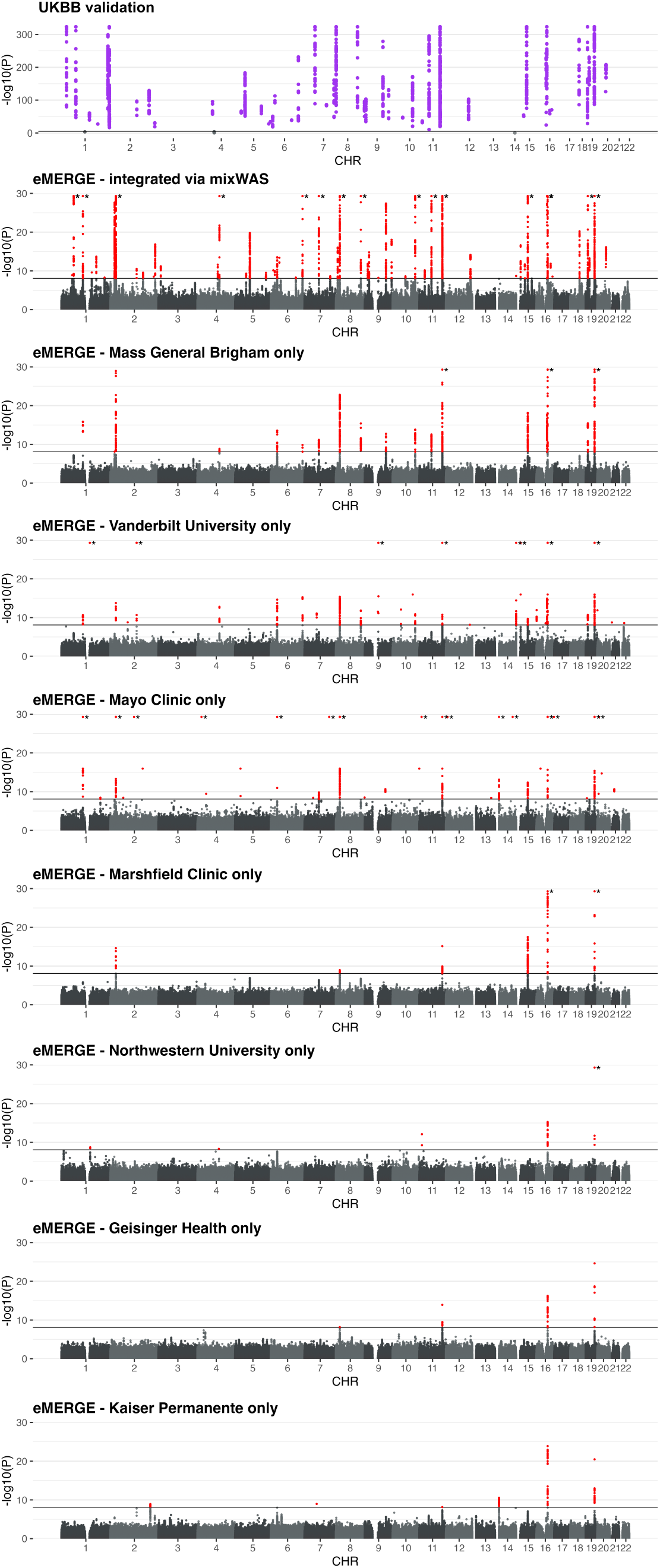
Manhattan plots of MPA in eMERGE single site analysis, eMERGE integrated analysis, and UKBB validation. The association between each SNP and all phenotypes was tested using mixWAS. The resulting p-value of each SNP was - log10(p) transformed and plotted along the y-axis. SNPs with p-values lower than 5e-30 were replaced with 5e-30 and indicated by asterisk for visualization purpose. The genomic position of the SNP was plotted along the x-axis. The solid horizontal line indicates Bonferroni corrected p-value significance level, respectively to each data. SNPs above the significance threshold are plotted as red points in the eMERGE data and purple points in the UKBB data.

### mixWAS identified MPA are associated with multiple traits/diseases

mixWAS-identified MPA could potentially be associated with one or multiple traits. To explore the specific trait/disease driving these MPAs, an exhaustive analysis evaluating all possible combinations of trait and SNP associations (4,530 SNPs x 10 traits) were conducted in the UKBB dataset. The significance of the single trait associations was determined using the mixWAS related p-value threshold (Method). The analysis revealed that SNPs showed significant associations with between 0 to 8 traits, with 2 to 4 traits being the most common number (Figure 4a).

**Figure 4.**
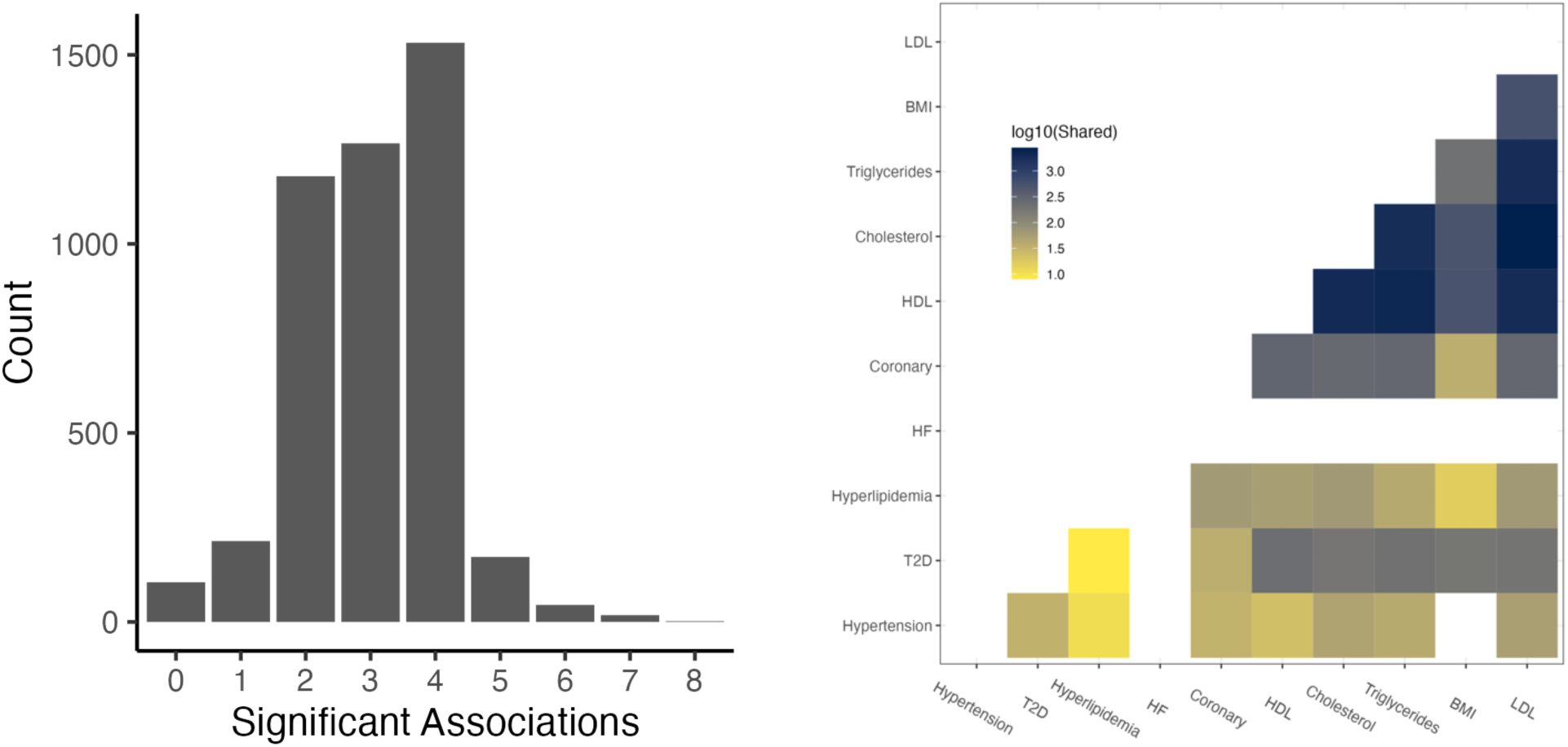
a) The distribution of the number of traits associated with a SNP. Count (y-axis) reflects the number of SNPs with significant associations for varying numbers of phenotypes (x-axis). Each association is determined using regression analysis, while adjusting for the Bonferroni-corrected p-value threshold. b) Common genetic variants shared among different traits. For each pair of phenotype combinations, the number of shared same SNP associations that surpassed the significant p-value threshold was calculated and transformed using a log10 scale. The color intensity in the plot reflects the number of shared genetic variants, with darker shades indicating a higher number of shared associations.

### Variability in shared genetic variants

The number of genetic associations common to different traits exhibited variability. Blood lipid levels (LDL, HDL, Cholesterol, and Triglycerides) and BMI displayed the highest number of shared genetic variants among the traits. Additionally, blood lipid levels demonstrated considerable overlap with genetic variants associated with coronary artery disease. For T2D, both blood lipid levels and BMI showed enrichment for shared genetic variants (Figure 4b).

### Improved detection powered by mixWAS

Among the initial pool of 4,530 candidate SNPs with detected MPAs across 10 traits, 13,770 significant single trait-SNP associations were identified in the UKBB dataset from. In comparison, directly applying PheWAS to the same SNPs in UKBB would only detect 11,581 significant associations due to the increased number of tests (Supplemental Table S2). Consequently, mixWAS detected 18.9% more trait-SNP associations in the UKBB dataset (Supplemental Figure S2). These additional associations were found for every trait except for heart failure, where neither mixWAS nor PheWAS identified any SNP associated with the disease (see Discussion). The results underscore the improved sensitivity and efficiency of mixWAS in detecting trait-SNP associations compared to traditional PheWAS approaches.

### Functional annotation of mixWAS SNPs

The mixWAS SNPs were annotated using the canonical pathways curated in the Human Molecular Signatures Database (MSigDB). These SNPs exhibited enrichments in pathways related to cholesterol metabolism, lipoprotein function, hyperlipidemia, as well as pathways associated with LDL, HDL, and triglycerides (Figure 5. These findings provide additional support for the genetic associations identified through mixWAS.

**Figure 5.**
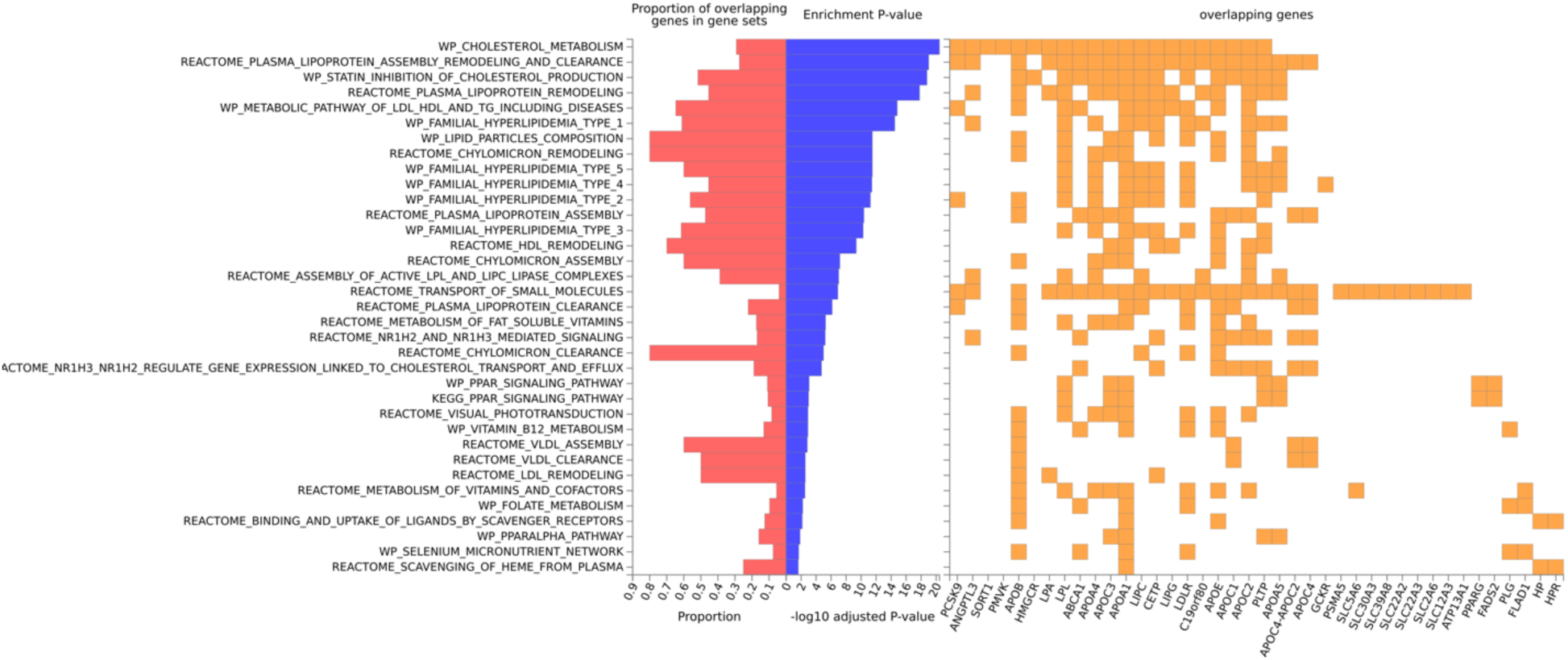
Pathway annotations of mixWAS identified SNPs. The mixWAS identified SNPs were annotated using the MSigDB canonical pathways. The enrichment of a pathway is determined by the number of SNPs associated with the pathway. The pathways are ranked by their enrichment -log10(p-values) from top to bottom (blue bar). SNPs were also mapped to genes within each pathway and their presence in a gene is represented by yellow squares. The proportion of overlapping genes in gene sets is shown as red bars.

## Discussion

Recent initiatives have made EHR-linked genetic data increasingly available for genomics research, providing extensive, well-characterized phenotype data that opens unprecedented opportunities for cross-cohort learning. The integration of genetic data with detailed clinical records from multiple health systems, institutions, and population studies allows for a more powerful and reproducible examination of genetic associations with multiple diseases and traits, illuminating potential shared genetic architectures among diverse phenotypes. However, despite these opportunities, significant data-sharing restrictions and methodological challenges limit the full potential of multi-EHR analyses in identifying multi-phenotype associations (MPA) and other cross-cohort genetic insights.

To overcome these barriers, we developed *mixWAS*, a computationally efficient, one-shot, lossless method designed for flexible integration of summary statistics across multiple datasets. This method enables robust identification of genetic variants associated with multiple phenotypes—both binary and continuous—and facilitates a comprehensive exploration of shared genetic variants underlying various traits. Beyond MPA detection, *mixWAS* is applicable to a wide range of multi-outcome studies across distributed data, underscoring its utility as a versatile tool for cross-cohort learning in complex multi-EHR settings.

We used simulation studies to demonstrate that mixWAS outperforms standard statistical approaches used in most PheWAS across a range of realistic settings that incorporate heterogeneity across sites, ranging direction, magnitude, and sparsity of phenotype effects, missing data, healthy volunteer biobanks, and common/rare genetic variants. By accounting for correlation between phenotypes in a manner that does not require individual level information, mixWAS gained the most power in settings where residual correlation existed between phenotypes, and SNP effects went against the correlation of these effects, a scenario often seen in real-world disease phenotypes^50^. In addition, type 1 errors were well controlled in all settings, as reflected by the power when the simulated effects equal to zero (Figure 2). Our simulations also demonstrate that methods which may work well for MPA detection in some settings, like multiPhen, don’t translate well to the federated setting when data sharing is not possible, and may breakdown even under moderate (10%) amounts of missing data.

Given that MPAs can often be difficult to detect, in large part due weak associations and multiple testing penalties in standard PheWAS methods, and given its improved power in most settings, mixWAS is a superior method for studying the shared genetic basis underlying multiple phenotypic traits in complex multi-EHR settings.

Towards this end, we employed the mixWAS method to study MPA across blood lipid levels, BMI, and diseases of the circulatory system using seven EHR sites from the eMERGE project, and we validated our findings using data from the UKBB. Supplemental Figure S1 illustrates heterogeneities in data characteristics across different eMERGE study sites. Notably, Vanderbilt and Mass General Brigham had the largest relative sample sizes compared to other sites, but both datasets had significant missing blood lipid measurements. The presence of differential missing data patterns is expected when integrating data from multiple real EHR sources, given the varying clinical protocols and patient populations among hospitals. Nevertheless, the mixWAS method can effectively account for the differential missing data across hospitals.

Applying mixWAS separately to each eMERGE site or across all sites yielded significantly different numbers of significant genetic associations. Comparing results between individual sites revealed a strong correlation between sample size and the number of detected genetic associations. Notably, the locations of the significant associations remained consistent between different datasets, suggesting the detection of the same MPAs across different EHRs, with only variations in the number of associations. The integrated eMERGE analysis identified the highest number of significant associations compared to any individual site (Figure 3). Importantly, the integrated analysis identified additional genetic associations that are not present in any single-site data, underscoring the benefits of this integrated approach.

The 4,530 mixWAS-identified MPA in eMERGE were further validated in the UKBB data. Using the p-value thresholds corresponding to the number of MPA, 4,428 MPA reached the significance threshold in UKBB (Figure 3). Given the distinct study populations and data generation processes between the two datasets (US and UK), we believe the 4,428 genetic variants represent robust MPAs for the studied diseases and traits. A common challenge in interpreting MPAs lies in distinguishing SNPs that are associated with only one phenotype from those associated with multiple phenotypes. However, a joint test, such as mixWAS, can effectively detect both types of associations equivalently. To further investigate the specific trait-SNP associations driving the MPAs, we performed additional single phenotype and SNP associations for all MPA SNPs identified in eMERGE. MPA SNPs were found to be significantly associated with 0 to 8 traits, with 2 to 4 traits being the most common, and the majority of MPA SNPs were associated with more than 2 phenotypes (Figure 4a).

Among the traits, lipid levels (including LDL, HDL, Cholesterol, and Triglycerides) shared the largest number of associated genetic variants, followed by BMI (Figure 4b). Additionally, coronary artery disease and T2D showed common MPAs with protein lipid levels and BMI. For heart failure, no significant associations were detected; however, some of the genetic associations were just below the significance threshold. The evaluated genetic variants are specifically those that showed MPAs across diseases, and the lack of identified associations may be due to limited number of heart failure cases, or shared genetic effects between heart failure and other diseases, or it may indicate inadequate power to detect smaller associations, or phenotype heterogeneity, or a combination of multiple factors. We additionally performed a separate GWAS analysis on heart failure alone in eMERGE and no SNP associations were found to be significant. This supports our hypothesis that the data was underpowered for studying heart failure. However, further studies are needed to confirm these results, as this study represents the initial identification of MPAs in these diseases and traits.

Moreover, we observed improved power in detecting specific trait-SNP associations from the 4,530 mixWAS-detected MPAs in UKBB. Compared to investigating all trait-SNP associations, or a PheWAS analysis, using mixWAS MPAs resulted in an 18.9% increase in the number of detected associations (Supplemental Figure S2). This increased number of associations can provide additional insights into the shared underlying genetics among different diseases and traits. Functional analysis of the mixWAS-detected MPA confirmed that the MPA are enriched for pathways related to cholesterol metabolism, lipoprotein function, hyperlipidemia, as well as pathways associated with LDL, HDL, and triglycerides (Figure 5). Together, these findings support that the mixWAS has improved power to detect more MPA that are functionally relevant to the studied diseases/traits.

In our evaluation of mixWAS, we tested it against a fixed number of phenotypes and found that it outperformed typical PheWAS statistical approaches. Importantly, this approach could be scaled to an arbitrarily large number of phenotypes, including different approaches taken to defining the phenome. mixWAS also extends its utility beyond genetic datasets and can be applied to any datasets that contain binary or continuous outcomes. Nevertheless, we also recognize several limitations of the study. First, while mixWAS can accommodate differential missing data patterns in each dataset, this relies on the assumption that the data is missing completely at random or missing at random. Second, the presence of a substantial number of null phenotypes—those unrelated to genetic variants—can diminish the power of mixWAS. Third, while mixWAS accounts for site-specific confounding effects, it primarily models linear effects averaged across sites. It does not directly capture individual-level confounding or complex non-linear interactions. However, it is possible to extend mixWAS to address shared non-linear confounders by first estimating site-specific non-linear effects and then sharing those estimates across sites. This represents a promising direction for future methodological development. Fourth, a key challenge in genetic research is the limited availability of large-scale, multi-ancestry genetic datasets linked to EHRs. While our current evaluation focused on datasets consisting primarily of individuals of European ancestry (eMERGE and UK Biobank), we acknowledge that genetic associations may differ across ancestral groups. While mixWAS is inherently ancestry-agnostic and applicable to multi-ancestry datasets, future studies are needed to validate its performance and assess the generalizability of MPAs across diverse populations as such data become more accessible. Lastly, mixWAS requires the sharing of more extensive summary-level statistics compared to traditional meta-analysis methods, resulting in higher data communication costs. However, these costs remain orders of magnitude lower than the transmission of entire datasets.

While the current study applies mixWAS to identify MPAs across genetically linked EHR datasets, the algorithm has broader applicability beyond genetic data. Distributed real-world datasets with mixed outcome types are common in settings such as clinically derived EHRs, drug response studies, and multi-omics research. The flexibility of mixWAS makes it well-suited for analyzing these types of data. Moreover, its applicability extends beyond biomedical domains—for example, to fields involving complex, distributed datasets with mixed data types, such as social sciences, economics, and environmental studies.

## Methods

### The proposed mixWAS Algorithm

mixWAS is designed to jointly test the association between a single SNP *X* and multiple mixed- type phenotypes (*Y*_1_, …, *Y_q_*), across *M* sites, which cannot share individual level patient data across sites due to privacy concerns. We let *i* index individuals, *j* index phenotypes, and *m* index sites. For the *i^th^* individual at the *m^th^* site we denote the SNP by *X_im_* ∈ {0, 1, 2 } and the *q* phenotypes by ***Y_im_*** = -*Y_i_*_1*m*_, …, *Y_iqm_* ∈ *R^q^*. The outcomes of interest may be of differing data types such as binary, continuous, count, or time to event.

Let ***Z***_***im***_ ∈ *R*\*denote a vector of an individual’s covariates, such as age, sex, or ancestry principal component (PCs). As a general framework for phenotype j,

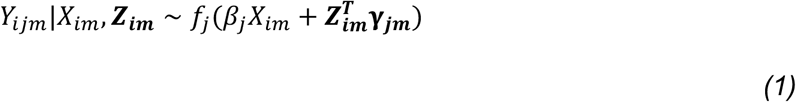

where β_+_ ∈ *R* denotes the SNP effect, shared across all *M* sites, and ***γ***_***jm***_ ∈ *R*\*denotes the site-, phenotype-specific effect sizes for remaining covariates. These effects are allowed to be site-specific to account for the site-level heterogeneity such as disease prevalence and confounding effects. Finally, *f_j_* denotes the density corresponding to the data type for each phenotype of interest. For example, binary outcomes can be assumed to follow the logistic regression model

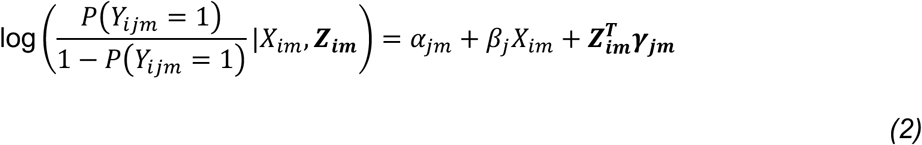

while continuous outcomes can be assumed to follow the linear regression model

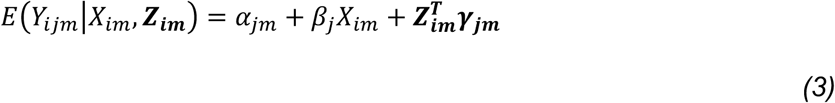

Following the approach of Li et al^1^, *q* phenotypes are combined using a composite likelihood function, which accounts for complex correlations between mixed-type phenotypes without modeling them directly^51,52^. The log composite likelihood function across all *M* sites can be expressed as

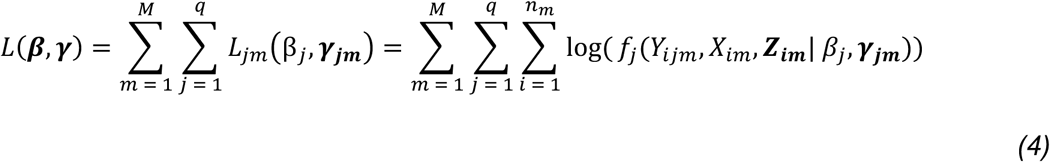

In order to determine whether a given SNP has association with any of the phenotypes of interest, we consider testing *H*_0_: 𝜷 = (𝛽_1_, …, 𝛽*_q_*) = **0** against *H*_1_: 𝜷 ≠ **0**, using an omnibus score-type test, which is computationally efficient and lossless in a federated setting in which sites cannot share individual level data. The score function is defined by

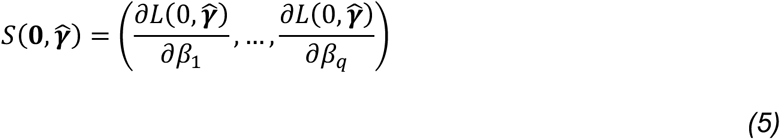

where ***γ̂*** are the maximum likelihood estimators of ***γ*** under *H*_2_ with 𝜷 set to 0. Non-SNP coefficients 𝛄_***jm***_ are both site- and phenotype-specific, and they can be estimated independently at each site by ***γ̂***_***jm***_ = argmax ***γ***_***jm***_ 𝐿*_jm_*-(**0**, 𝛄_***jm***_). Because of the composite likelihood framework, the score function can be decomposed into a sum of site-specific score vectors as follows

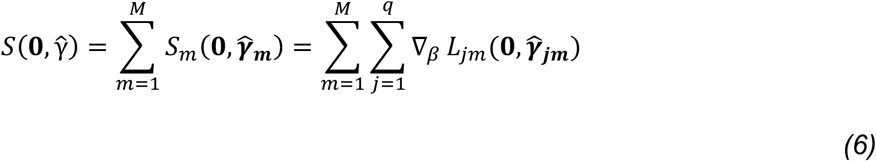

Thus, each site only needs to compute and share its own score vector *S_m_*(**0**, ***γ̂***_𝒎_), and associated *q* × *q* covariance matrix *V_m_*, which contains only summary-level information. *V_m_* can be estimated locally by deriving the influence functions.

Figure 1 outlines the full mixWAS algorithm, while Algorithm S1 in the supplementary material provides pseudo-code. mixWAS is lossless in the sense that there is no approximation error brought by the federated decomposition and we obtain identical results as the pooled analysis. It is also communication-efficient because only one round of communication is required across sites. Also, it is highly computationally efficient since the reduced model is shared across all genetic variants and the summary-level statistics have closed-form expressions.

### Comparisons with PheWAS

PheWAS methods were utilized as a set of baselines against which to compare mixWAS. For continuous phenotypes, a linear regression was fit for each phenotype, while logistic regressions were fit for each binary phenotype. We first consider site specific estimates 𝛽̂*_jm_* for phenotype *j* at site *m*, as sites can not pool their individual data, and combine estimates across sites using inverse-variance weighting to obtain -𝛽_1_, …, 𝛽̂*_q_*1. The overall *p*-value for the SNP is obtained in three ways: 1) by taking the minimum of the *q* Bonferonni adjusted *p*-values (min P); 2) by applying HMP to the *q p*-values; 3) by applying ACAT to *q p*-values.

### Federated MultiPhen

MultiPhen was also used as a comparator for mixWAS. At each site, multiPhen was run on the *q* phenotypes, producing a *p*-value for the joint model at each site (e.g., for the model testing all *q* phenotypes simultaneously)^53^. The final *p*-value for the SNP is obtained by taking the minimum of *M* Bonferonni adjusted *p*-values.

### Simulating MPA across multiple EHRs

Various multi-phenotype association models were simulated to compare mixWAS with existing baselines (Supplemental Materials). As mixWAS is designed to integrate summary-level data across sites when individual-level data is unable to be pooled, we generate data at 5 sites. For comparison against mixWAS, we compute three PheWAS and MultPhen methods, as described in the sections above.

Methods were additionally compared to an oracle score test, which is a score test using only de- correlated 𝑧 −scores for non-null phenotypes. This test is considered an oracle test because in which phenotypes have non-null associations with the SNP is unknown in practice. As such, this reference gives a helpful upper bound on the power of score-based hypothesis tests under this setting.

Power curves from the simulation are shown in Figure 2. Most notably, mixWAS has higher power compared to standard PheWAS methods in nearly all scenarios, especially in cases where only 2 of the 8 SNP effects are non-null, and when the SNP effects oppose the direction of residual correlation between phenotypes.

### Utilizing mixWAS to detect MPA using eMERGE and UK Biobank

From the eMERGE study (Supplemental Material), patients from multiple adult electronic health records (EHRs), including Marshfield Clinic, Vanderbilt University, Kaiser Permanente/University of Washington, Mayo Clinic, Northwestern University, Geisinger, and Mass General Brigham, were included in the research. Binary disease statuses for individuals were determined based on specific ICD-9 codes, including unspecified essential hypertension (ICD-9 401.9), type 2 diabetes (ICD-9 250.00), hyperlipidemia (ICD-9 272.4), benign essential hypertension (ICD-9 401.1), atrial fibrillation (ICD-9 427.31), congestive heart failure (ICD-9 428.0), and coronary atherosclerosis (ICD-9 414.00). Additionally, median laboratory measures, including LDL, HDL, serum total cholesterol, triglycerides, and BMI for each patient, were calculated and utilized as continuous outcomes.

mixWAS was applied to each SNP to detect MPA among the mixed binary and continuous outcomes. This analysis included adjustments for age, sex, and the top 10 principal components to account for population stratification. Given the different ages of disease onset for each condition, distinct disease-associated ages were incorporated into the mixWAS model. The disease-associated age for each individual was computed as the median age for each continuous laboratory measure, corresponding to the median laboratory measures used for the analysis For binary diseases, the median age of the ICD-9 code assignments for each individual of a disease was employed as the age for cases, while for controls, the age was determined as the patients’ age at their last EHR record.

The MPA identified by eMERGE were independently validated using data from the UK Biobank (Supplemental Material). Since the UKBB primarily utilizes ICD-10 codes for clinical diagnosis, a mapping process was carried out to convert the ICD-9 codes used in eMERGE data to their corresponding ICD-10 codes. The converted ICD-10 codes were unspecified essential hypertension and benign essential hypertension (ICD-10 I10), type 2 diabetes (ICD-10 E119), hyperlipidemia (ICD-10 E784 and ICD-10 E785), atrial fibrillation (ICD-10 I489), congestive heart failure (ICD-10 I509), and coronary atherosclerosis (ICD-10 I251). The continuous laboratory measures were extracted from the following fields, including LDL (field 30780), HDL (field 30760), total cholesterol (field 30690), triglycerides (field 30870), and BMI (field 12001). Notably, unspecified essential hypertension (ICD-9 401.9) and benign essential hypertension (ICD-9 401.1) from eMERGE were consolidated into a single condition, essential (primary) hypertension (ICD- 10 I10), in the UKBB dataset.

In the eMERGE discovery analysis, the significance threshold for SNPs’ p-value was set as 8.19 x 10^-9^ (0.05/6,106,952), corresponding to the Bonferroni adjusted p-value threshold. Subsequently, the 4,530 significant SNPs identified were re-evaluated in the UKBB dataset using the mixWAS algorithm, with a significance threshold set at 0.05/4,530=1.103 x 10^-5^. Furthermore, these 4,530 significant SNPs underwent PheWAS analysis in the UKBB to identify specific SNP-phenotype associations driving the MPAs. The Bonferroni-adjusted p-value threshold for this analysis was set at (0.05/4,530)/10 = 1.103 x 10^^-6^. In contrast, the standard PheWAS Bonferroni-corrected p-value threshold is 7.90 x 10^^-10^, which accounts for analyzing all SNPs in the UKBB dataset.

Functional annotation of the mixWAS-identified SNPs was carried out using the FUMA software^54,55^, and these SNPs were annotated using canonical pathways from the Human Molecular Signatures Database (MSigDB)^56^.

## Data Availability

UK Biobank and eMERGE

## Resource availability

### Lead contact

Requests for resources should be directed to the lead contact, Ruowang Li

### Materials availability

This study did not generate new materials.

### Data and code availability

The mixWAS algorithm and the code associated with this study have been deposited at: https://github.com/lbenz730/mixWAS as well as on Figshare: https://doi.org/10.6084/m9.figshare.28873547.v257

## Acknowledgment

NIH R01 LM010098, AG066833, GM148494, LM014344, LM012607, LM013519, AI130460, AG073435, RF1AG077820, R56AG069880, R56AG074604, U01TR003709, R21AI167418 and R21EY034179. MDR was funded by R01HG010067 and R01HL169458.

eMERGE Network (Phase III). This phase of the eMERGE Network was initiated and funded by the NHGRI through the following grants: U01HG8657 (Group Health Cooperative/University of Washington); U01HG8685 (Brigham and Women’s Hospital); U01HG8672 (Vanderbilt University Medical Center); U01HG8666 (Cincinnati Children’s Hospital Medical Center); U01HG6379 (Mayo Clinic); U01HG8679 (Geisinger Clinic); U01HG8680 (Columbia University Health Sciences); U01HG8684 (Children’s Hospital of Philadelphia); U01HG8673 (Northwestern University); U01HG8701 (Vanderbilt University Medical Center serving as the Coordinating Center); U01HG8676 (Partners Healthcare/Broad Institute); and U01HG8664 (Baylor College of Medicine).

UK Biobank. All data for this cohort pertained to project 32133 – “Integration of multi-organ imaging phenotypes, clinical phenotypes, and genomic data”.

## Author Contributions

R.L., L.B., R.D., J.M., Y.C., designed and conducted the study. R.L., J.M., Y.C., supervised the study. All authors contributed to the formation of the manuscript.

