## Supplementary Material for "A One-Shot Lossless Algorithm for Cross-Cohort Learning in Mixed-Outcomes Analysis"

**Supplementary Figures**

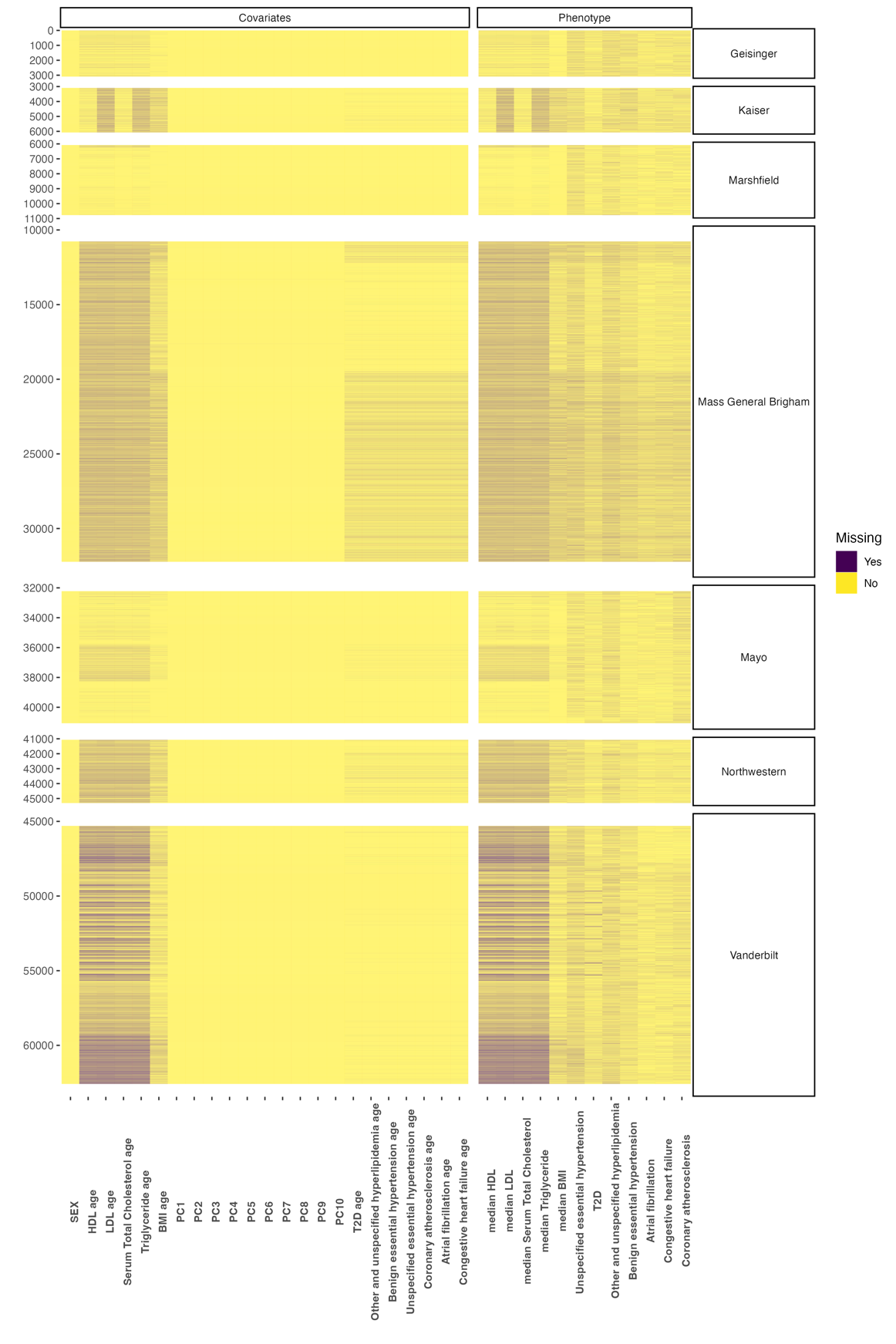
**Figure S1.** Characteristics of the covariates and phenotypes from the eMERGE dataset. The x-axis displays the phenotypes and covariates, and y-axis displays the number of participants in each dataset. The color indicates whether or not a value (binary or continuous) was present for the individual.

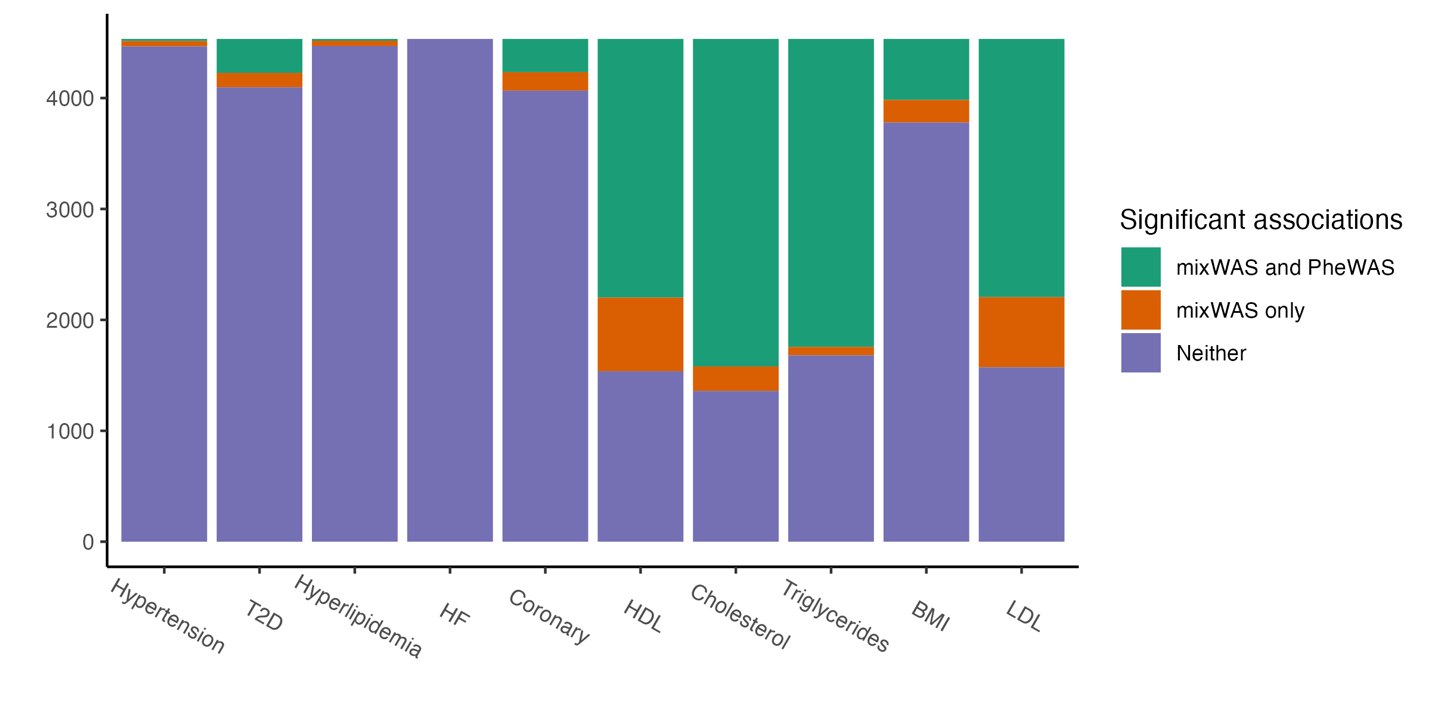

**Figure S2. Count of significant trait-SNP associations identified by mixWAS and PheWAS in the UKBB.** Method-specific p-value thresholds were applied to mixWAS and PheWAS, corresponding to the number of comparisons performed. The associations detected by PheWAS are a subset of those identified by mixWAS, and are categorized as "mixWAS and PheWAS”. The "mixWAS only" category represents genetic associations that were exclusively identified by mixWAS and not by PheWAS. SNPs that were not considered significant by either method are represented by the "Neither" category.

**EHR datasets**

**eMERGE data**

Genotype data linked with EHR information from the eMERGE network Phase III. This dataset comprised a total of 83,717 genotyped patients across 11 participating sites^40^. For our investigation, we focused on eight adult sites, which included the Marshfield Clinic Research Foundation, Vanderbilt University Medical Center, Kaiser Permanente Washington/University of Washington, Mayo Clinic, Northwestern University, Geisinger, Mt. Sinai, and Mass General Brigham. SNPs were imputed using the Haplotype Reference Consortium 1.1 reference, aligned with genome build 37. This imputation process yielded a set of 39 million genetic variants^41^. Subsequently, we subjected the SNP genotypes to quality filtering and processing, following an established pipeline^42^. The criteria for inclusion required that both the genotype and sample call rates exceeded or equaled 99%, the imputation score exceeded 0.4, Hardy-Weinberg equilibrium p-value used was 0.00001, and the Minor Allele Frequency (MAF) of the SNPs was equal to or exceeded 0.05. To mitigate the potential impact of population structure on our analyses, we restricted our investigation to unrelated individuals of European ancestry. The ancestry was determined using principal components derived from the 1000 Genome Project. In cases where individuals were identified as related, defined by a π-hat value of ≥ 0.25 identity-by-descent, one individual from each related pair was removed. The final dataset comprised 59,136 unrelated individuals, and a curated set of 6,106,952 high-quality SNPs for subsequent analyses.

**UK BioBank data**

The UK Biobank released comprehensive genetic and phenotypic data, encompassing approximately 500,000 individuals representing diverse regions across the United Kingdom^43^. Genotyping was conducted utilizing two related types of genotype arrays, namely the UK BiLEVE Axiom Array or the UK Biobank Axiom Array, organized into 106 batches and imputed using the merged UK10K and 1000 Genomes phase 3 reference panels^44^.

To ensure the quality and reliability of our sample, a series of quality control measures were implemented. First, individuals displaying a SNP missing rate exceeding 5% and exhibiting high levels of heterozygosity were excluded from the study. Second, among related individuals, one individual from each pair was systematically removed to prevent undue influence from familial genetic connections. The threshold for relatedness was set at the level of second-degree relatives, as indicated by an identity-by-descent π-hat value equal to or greater than 0.25. Third, only individuals with White British ancestry were retained in order to match the ancestry of the eMERGE data. Additionally, individuals with discrepancies between their self-reported and genetically-inferred sexes were omitted from the analysis. Finally, genetic variants characterized by imputation info scores lower than 0.3 and MAF less than 0.01 were excluded from consideration.

**Account for missing data and sparse alternatives**

mixWAS does not require all $M$ sites to have collected data on all $q$ phenotypes. Additionally, mixWAS does not require all individuals to have data available for all phenotypes collected at the site, provided any missing phenotype data at site level is missing completely at random (MCAR) or missing at random (MAR) in a manner described by covariates $\boldsymbol{Z}_{\boldsymbol{im}}$**.** Specifically, let $\delta_{ijm}$ be an indicator denoting whether $Y_{ijm}$ is observed. Then the composite likelihood in equation (4) can be modified as

$$L\left( \boldsymbol{\beta},\boldsymbol{\gamma} \right)=\sum_{m = 1}^{M} \sum_{j = 1}^{q} L_{jm}\left( \beta_{j},\boldsymbol{\gamma}_{\boldsymbol{jm}} \right)=\sum_{m = 1}^{M} \sum_{j = 1}^{q} \sum_{i = 1}^{n_{m}} \delta_{ijm} \log(f_{j}(Y_{ijm},X_{im},\boldsymbol{Z}_{\boldsymbol{im}}\boldsymbol{|}\beta_{j},\boldsymbol{\gamma}_{\boldsymbol{jm}}\boldsymbol{)})$$

(S1)

Note that when a site has no data on a particular phenotype collected, its contribution to the entry of overall score vector corresponding to that phenotype is just 0. We can obtain an overall test of $H_{0}:\boldsymbol{\beta}=\boldsymbol{0}$ by

$$T=S^{T}V^{-1}S=\left( \sum_{m=1}^{M} S_{m}\left( 0,{\hat{\boldsymbol{\gamma}}}_{\boldsymbol{m}} \right) \right)^{T}\left( \sum_{m=1}^{M} V_{m} \right)^{-1}\left( \sum_{m=1}^{M} S_{m}\left( 0,{\hat{\boldsymbol{\gamma}}}_{m} \right) \right)$$

(S2)

Asymptotically, $T\sim\chi_{q}^{2}$, which we can leverage to obtain a corresponding $p$-value, $p_{score}$. This test does well for dense alternatives when many of the $q$ phenotypes are non-null ($\beta_{j}\neq0$ for many of the $j \in\{1, \ldots, q\}$). However, when $q$ is large and most phenotypes are not significantly associated with the SNP (for example only one or two $\beta_{j}$ are non-zero), this score test may not be sufficiently powerful as the signal could be diluted by the majority of null effects.

Thus, we also consider a second test that is powerful under sparse alternatives, when the majority of the SNP effects are zero. Specifically, let

$$\boldsymbol{z}=V^{-1/2}S = \left( \sum_{i=1}^{m} V_{m} \right)^{-1/2}\left( \sum_{i=1}^{m} S\left( \boldsymbol{0},{\hat{\boldsymbol{\gamma}}}_{\boldsymbol{m}} \right) \right)$$

(S3)

$\boldsymbol{z}=\left( z_{1},\ldots,z_{q} \right)\sim^{\mathrm{iid}}N\left( 0,1 \right)$ and thus $p$-values $p_{1},\ldots,p_{q}$for the corresponding hypotheses $H_{0}:\beta_{j}=0$ can be obtained $p_{j}=2\Phi\left( -\left| z_{j} \right| \right)$ where $\Phi\left( \cdot\right)$ is the CDF of the standard Normal distribution. In order to combine the $p$-values, we use the aggregated Cauchy association test (ACAT)^47^.

$$t_{ACAT}=\frac{1}{q}\sum_{j=1}^{q} \tan\left( \pi\left[ \frac{1}{2}-p_{j} \right] \right)$$

$$p_{ACAT}=\frac{1}{2}-\frac{1}{\pi}\tan^{-1} \left( t_{ACAT} \right)$$

(S4)

Originally developed as a fast, computationally efficient $p$-value combination method for rare variant analyses, ACAT was shown particularly powerful in the presence of only a small number of causal variants in a variant set. Via simulations, we will show that such a test boosts power in the case of sparse alternatives.

ACAT has also been shown to be useful as a method for combining p-values from tests powerful in differing scenarios to create an omnibus test^47^. It is particularly appealing as a way to combine our two component $p$-values, as it does not require one to estimate or account for potentially very complex correlation between component $p$-value. Thus, in order to create a test robust to both dense and sparse alternatives, we use ACAT to combine $p_{score}$, which is powerful against dense alternatives, and $p_{ACAT}$, which boosts power against sparse alternatives, as shown in Equation (S5).

$$t_{SNP}=\frac{1}{2}\left( \tan\left( \pi\left[ \frac{1}{2}-p_{score} \right] \right)+\tan\left( \pi\left[ \frac{1}{2}-p_{ACAT} \right] \right) \right)$$

$$p_{SNP}=\frac{1}{2}-\frac{1}{\pi}\tan^{-1} \left( t_{SNP} \right)$$

(S5)

**Additional details on simulating MPA across multiple EHRs and additional simulation settings**

We first outline a general process for generating data in simulations. We begin by drawing six covariates for each subject at each of $M=$ 5 sites, denoted by $\boldsymbol{Z}_{\boldsymbol{im}}\boldsymbol{=}$ 4 principal components (PCs), age, and sex. Corresponding coefficients for these covariates*,* $\boldsymbol{\gamma}_{\boldsymbol{jm}}$ that are both site- and phenotype-specific are generated for each phenotype and site. Distributional choices for generating $\boldsymbol{Z}_{\boldsymbol{im}}$and $\boldsymbol{\gamma}_{\boldsymbol{jm}}$ are shown in Table S4.

| **Variable** | $\boldsymbol{Z}_{\boldsymbol{im}}$ **Generation** | $\boldsymbol{\gamma}_{\boldsymbol{jm}}$ **Generation** |
| --- | --- | --- |
| Principal Components (4) | $N(0, 1)$ | $Uniform(-0.5, 0.5)$ |
| Age (Centered) | $N\left( 0,{15}^{2} \right)$ | $Uniform(-0.05, 0.05)$ |
| Sex | $Bernoulli(0.5)$ | $Uniform(-0.1, 0.1)$ |

**Table S4**: Data generation mechanism for individual covariates ($\boldsymbol{Z}_{\boldsymbol{im}}$) and corresponding coefficients ($\boldsymbol{\gamma}_{\boldsymbol{jm}}$). Note that $\boldsymbol{\gamma}_{\boldsymbol{jm}}$ coefficient vectors are generated for each phenotype independently.

SNPs $X$ are drawn from $\text{Binomial}\left( 2, \text{MAF} \right)$, where the minor allele frequency (MAF) is a parameter of the specific simulations. SNPs are centered to have mean 0 by subtracting $E\left[ X \right]=2\times\text{MAF }$so that the size of any SNP effect does not change the prevalence of any binary phenotype. $q_{c}$ continuous phenotypes are generated from the following linear model

$$Y_{ijm}=\beta_{j}X_{im}+\boldsymbol{Z}_{\boldsymbol{im}}^{\boldsymbol{T}}\boldsymbol{\gamma}_{\boldsymbol{jm}}+\epsilon_{ijm}$$

(S6)

Random noise $\boldsymbol{\epsilon}_{\boldsymbol{im}}=\left( \epsilon_{i1m,}\ldots,\epsilon_{iq_{c}m} \right)\sim N\left( \boldsymbol{0},\boldsymbol{\Sigma}_{\boldsymbol{c}} \right)$is added to each subjects’ phenotype vector $\boldsymbol{Y}_{\boldsymbol{im}}$ to induce covariance $\boldsymbol{\Sigma}_{\boldsymbol{c}}$ across phenotypes. $q_{b}=q-q_{c}$ binary phenotypes are generated by the following logistic regression model

$$\log\left( \frac{P\left( Y_{ijm}=1 \right)}{1-P\left( Y_{ijm}=1 \right)} |X_{im},\boldsymbol{Z}_{\boldsymbol{im}} \right)=\alpha+\beta_{j}X_{im}+\boldsymbol{Z}_{\boldsymbol{im}}^{\boldsymbol{T}}\boldsymbol{\gamma}_{\boldsymbol{jm}}$$

(S7)

where $\alpha=\log\left( \frac{\text{Prevalence}}{1-\text{Prevalence}} \right)$ and the prevalence = $E\left( Y_{ijm} \right)$, which is a simulation parameter, is set to be the same for each binary phenotype. Each subjects $q_{b}$ binary phenotypes are drawn using the rmvbin() in R’s bindata library^48^, which creates correlated multivariate binary random variables by thresholding a normal distribution. Marginal probabilities are obtained by applying the inverse logit function to equation (S7), and covariance matrix $\boldsymbol{\Sigma}_{\boldsymbol{b}}$ is used to induce a correlation for the resulting binary phenotypes. Let $\boldsymbol{\Sigma}=\left( \begin{matrix} \boldsymbol{\Sigma}_{\boldsymbol{c}} & \boldsymbol{0} \\ \boldsymbol{0} & \boldsymbol{\Sigma}_{\boldsymbol{b}} \end{matrix} \right)$ denote the overall covariance matrix for all phenotypes. In all simulations, 10% of phenotypes are set to be missing completely at random, and additionally we do not require that each site has all data available for all $q$ phenotypes.

We consider $\boldsymbol{\beta}=\left( \beta_{1},\ldots,\beta_{q} \right)$ of varying direction and sparsity. We define the sparsity of $\boldsymbol{\beta}$ to be the number of components that are non-zero. All simulations set $q = 8$, and we consider sparsities ranging from $\beta=\left( \beta,\beta,0,0,0,0,0,0 \right)$ (2 non-null phenotypes) to $\boldsymbol{\beta}=\left( \beta,\beta,\beta,\beta,\beta,\beta,\beta,\beta\right)$ (8 non-null phentypes). While we always set the magnitude $\left| \beta_{j} \right|= \beta$ to be the same for each non-null phenotype, we consider cases where all non-zero components are positive (e.g. $\boldsymbol{\beta}=\left( \beta,\beta,\beta,\beta,0,0,0,0 \right)$), all non-zero components are negative (e.g. $\boldsymbol{\beta}=\left( -\beta,-\beta,-\beta,-\beta,0,0,0,0 \right)$*),* and a case where the direction of the effects are opposite, with half non-zero components being positive and half being negative (e.g. $\boldsymbol{\beta}=\left( \beta,-\beta,\beta,-\beta,0,0,0,0 \right)$). For each magnitude of effect, $\beta$, 2000 simulated datasets were generated, and power for each method was calculated as the percentage of times a method identified significant multi-phenotype associations out of 2000 repetitions.

To demonstrate the utility of mixWAS, we first consider a simulation setting in which $q$= 8 phenotypes are available at $M$ = 5 sites, with $n_{m}$ = 1000 subjects at each site. Of the 8 available phenotypes, $q_{c}$ = 4 are continuous $\left( \boldsymbol{Y}_{\boldsymbol{1}}\boldsymbol{,\ldots,}\boldsymbol{Y}_{\boldsymbol{4}} \right)$ and $q_{b}$ = 4 are binary $\left( \boldsymbol{Y}_{\boldsymbol{5}}\boldsymbol{,\ldots}\boldsymbol{Y}_{\boldsymbol{8}} \right)$. A common variant setting was chosen for binary phenotypes with MAF = 20% and the prevalence of each phenotype = 30%. Effect sizes $\left| \beta\right|$ considered ranged from [0.01, 0.35].

For this setting, binary phenotypes are always considered to be positive, indicating a SNP increases the likelihood of each phenotype (disease). We consider cases where 2 (1 binary + 1 continuous), 5 (3 binary + 2 continuous), and all 8 $\beta_{j}$ are non-zero. While binary phenotypes are always positive, we consider cases where the continuous phenotypes either all positive (Figure 2A) or are in opposite directions (e.g. 1 negative, 1 negative/1 positive, and 2 negative/2 positive in the 3 levels of sparsity, respectively) [Figure 2B].

We consider 3 types of residual correlation between phenotypes: positive, no correlation, and negative. Correlations between phenotypes are shown in Figure S3. When all effects are positive, residual correlation is added to both continuous and binary phenotypes. In the case of opposite direction continuous phenotypes, we only include residual correlation for continuous phenotypes. As this setting only considers positive binary phenotype, we wanted to understand the interaction between oppositive direction effects and different directions of correlations without power varying due to the direction of residual correlation of the positive binary effects, which can heavily influence power (Figure 2). Note that $\boldsymbol{\Sigma}_{\boldsymbol{c}}$, the covariance matrix for the continuous phenotypes is simply the upper block diagonal of the correlation matrix scaled by $\sigma^{2}={2.3}^{2}$.

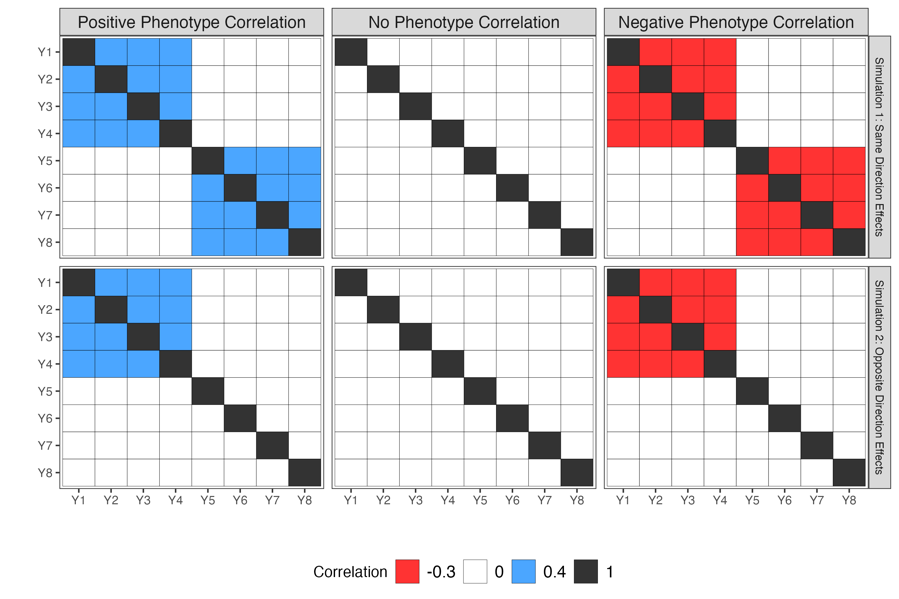

**Figure S3:** Correlation matrix for mixed data type simulation. $\left( \boldsymbol{Y}_{\boldsymbol{1}},\ldots,\boldsymbol{Y}_{\boldsymbol{4}} \right)$ are continuous phenotypes while $\left( \boldsymbol{Y}_{\boldsymbol{5}},\ldots\boldsymbol{Y}_{\boldsymbol{8}} \right)$ are binary phenotypes. To get the full covariance matrix for the continuous phenotypes, $\boldsymbol{\Sigma}_{\boldsymbol{c}}$, the upper block diagonal is scaled by $\sigma^{2}={2.3}^{2}$. For binary phenotypes, the correlation matrix is supplied to R function rmvbin() in the bindata package.

**Binary Phenotypes: Common and Rare Variant Settings**

In the additional simulation study, we also consider a method that is more common in pleiotropy analysis, ASSET^48^, which exhaustively searches subsets of the phenotypes for significant effects, and is able to account for correlation between the phenotypes introduced by sample overlap. ^43^. Since ASSET is only for pleiotropic analyses of binary phenotypes, we consider various simulation settings in which all 8 phenotypes $\left( \boldsymbol{Y}_{\boldsymbol{1}}\boldsymbol{,\ldots,}\boldsymbol{Y}_{\boldsymbol{8}} \right)$ are binary. As outlined above, $q=q_{b}=8,n_{m}=1000$ subjects at each of $M=5$ sites. We consider a common variant setting with MAF = 20% and prevalence = 30%, along with a rare variant setting with MAF = 5% and prevalence = 10%. Effect sizes $\left| \beta\right|$ considered ranged from [0.01, 0.60]. $\boldsymbol{\Sigma=}\boldsymbol{\Sigma}_{\boldsymbol{b}}$ is shown in Figure S7a. Only positive correlation is considered in this setting.

Empirical power curves for this pair of binary phenotype simulations are shown in Figure S4. In the common variant/high prevalence setting, mixWAS outperforms all PheWAS methods, including ASSET, which is overly conservative in most settings, as it only accounts for correlation due to sample overlap, rather than correlation induced by the fact that outcomes themselves may be highly correlated across subjects. In the rare variant/low prevalence setting, PheWAS methods are highly impacted by the direction of non-null effects. When effects are all positive (i.e. SNP increases disease prevalence), PheWAS outperforms even the oracle score test.

However, when all SNP effects are negative (SNP decreases disease prevalence) PheWAS does significantly worse than mixWAS, due to the fact that the direction of these effects is against the direction of the residual correlation (Figure S4a). Similar results can be seen when effects are in opposite directions. The sign of the effect likely impacts PheWAS methods in low prevalence settings due to the fact that logistic regression is biased in problems with heavy class imbalance^49^

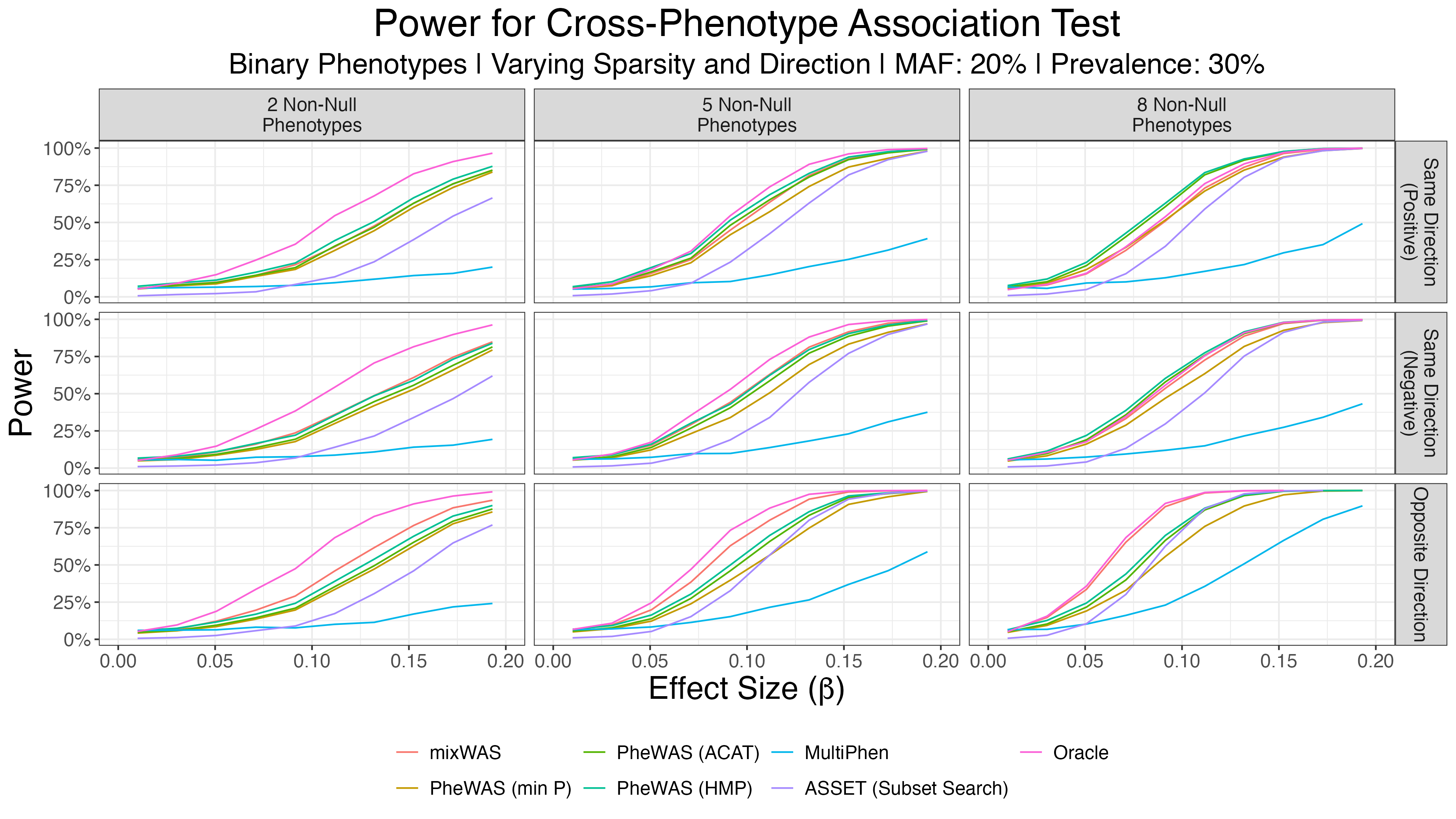
**a:** Common Variant/High Prevalence Setting

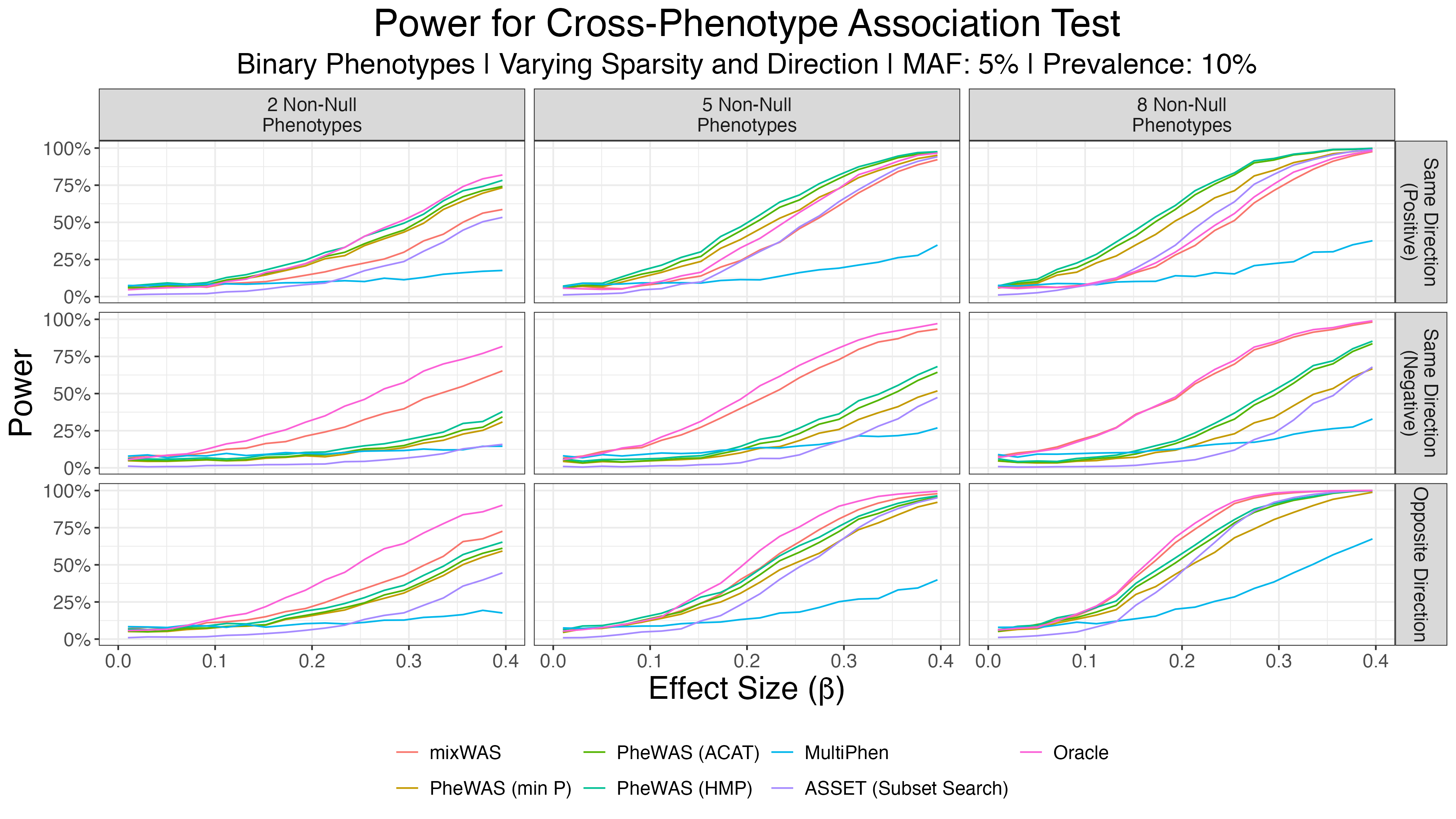

**b:** Rare Variant/Low Prevalence Setting

**Figure S4:** Empirical power curves for binary phenotype simulations. In the common variant/high prevalence setting, mixWAS outperforms all PheWAS methods, including ASSET, which is overly conservative in most settings. In the rare variant/low prevalence setting, PheWAS methods are highly impacted by the effect direction(s), doing better than mixWAS in the case of all positive effects but performing significantly worse when all effects are negative, or effects are in opposite directions.

**Healthy Controls**

Genetic biobanks, like the UK Biobank, often only contain controls with none of the diseases/phenotypes of interest, and cases for each specific subject. A subject may be a case for multiple phenotypes, but the absence of a subject being a case for a particular phenotype does not make them a control for that phenotype–their disease status is simply unknown, and no subject can ever be both a case one phenotype and a control for a different phenotype.

To mimic the structure of these real-life genetic databases, we designed a simulation with healthy controls. 8 binary phenotypes were generated in the same manner as the common variant, high prevalence binary phenotypes above, except that controls were designated as subjects for whom $Y_{ijm}$= 0 for $j \in\{1, \ldots, 8\}$. Subjects who were a case for at least one phenotype had their control status for other phenotypes replaced by an *NA* indicating disease status unknown. A stronger correlation structure $\boldsymbol{\Sigma=}\boldsymbol{\Sigma}_{\boldsymbol{b}}$was utilized in this simulation, as shown in Figure S7b. Note that due to the perfect split between cases and controls, an even stronger correlation is induced between phenotypes than what is shown in Figure S6b. Additionally, since subjects who were cases for some subset of phenotypes could be controls for other phenotypes in previous simulations, but under the healthy control setting cannot be controls for other phenotypes, the effective number of subjects is drastically reduced compared to other simulation settings.

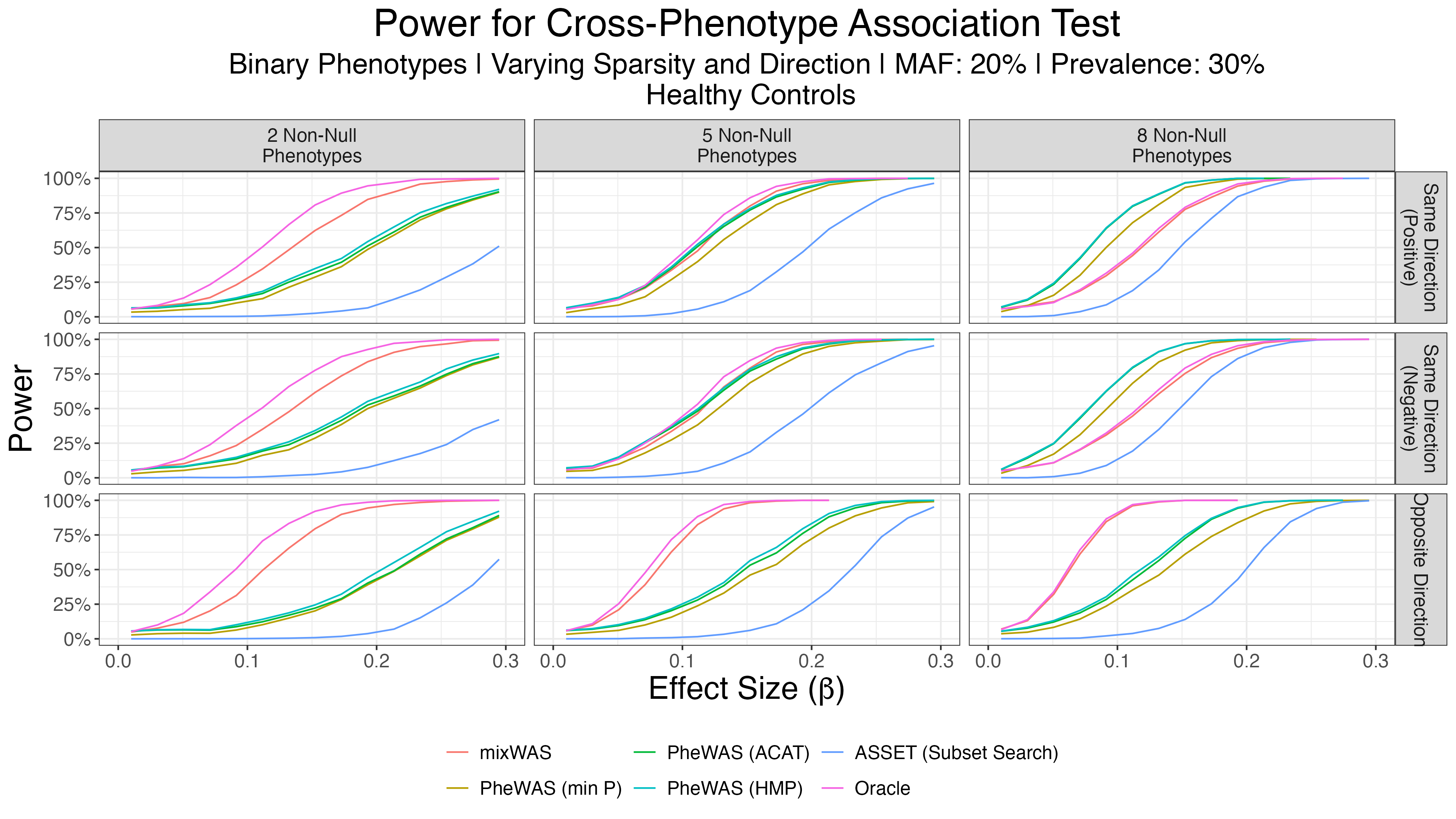

**Figure S5:** Empirical power curves comparing various cross-phenotype association tests for simulated binary phenotypes using only healthy controls to compare against diseased cases. mixWAS significantly outperforms ASSET in every setting, as the latter method does not account for correlation between phenotypes except for correlation induced by sample overlap. PheWAS obtains the highest power when no-sparsity is present, but power is again affected by the direction of effects.

Empirical power curves are shown in Figure S5. mixWAS significantly outperforms ASSET in every setting, as the latter method does not account for correlation between phenotypes except for correlation induced by sample overlap. PheWAS obtains the highest power when no sparsity is present, but power is again affected by the direction of effects, with power being worst under mixed-direction phenotypes. This result is likely explained once again by the smaller effective sample size introduced by limiting our previous simulated sample to include only health controls as well as the bias of logistic regression in class imbalanced problems^49^.

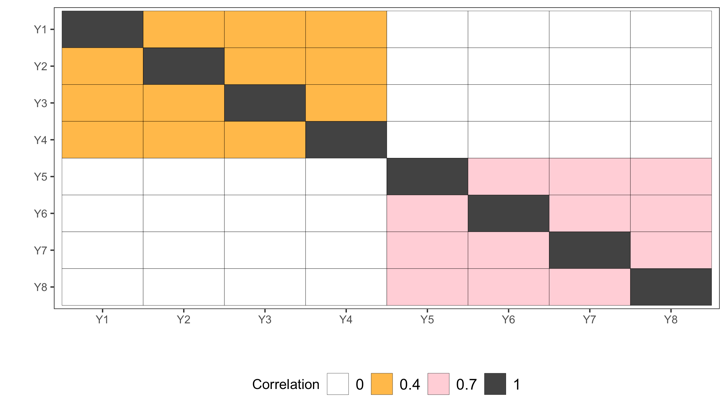

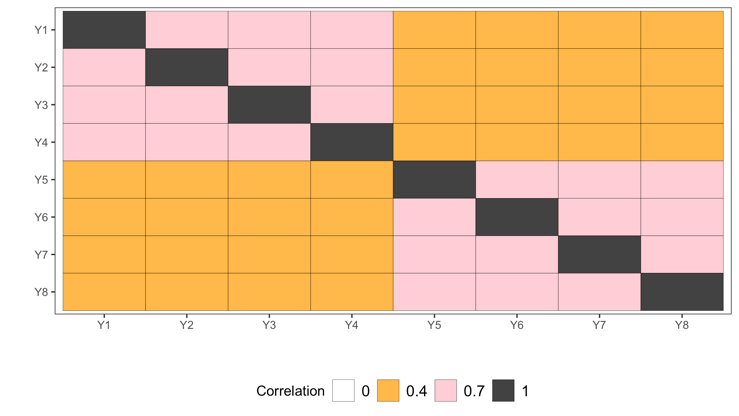

**a:** Common and Rare Variant Settings	**b**: Health Controls Setting

**Figure S6**: Correlation matrices for binary phenotype data type, supplied to R function rmvbin() in the bindata package^44^.

**Simulation Under Differential Missing Mechanism**

To verify that mixWAS performed well under missing at random (MAR) outcomes, the main simulation setting (Figure 2A) was re-run with phenotypes missing at random rather than completely at random (MCAR). In particular the following logistic regression was used to simulate missingness.

$$\log\left( \frac{P\left( \delta_{ijm}=1 \right)}{1-P\left( \delta_{ijm}=1 \right)} |X_{im},\boldsymbol{Z}_{\boldsymbol{im}} \right)=\zeta+\boldsymbol{Z}_{\boldsymbol{im}}^{\boldsymbol{T}}\boldsymbol{\eta}_{\boldsymbol{jm}}$$

For this setting, $\zeta=logit($0.08), $Z$ was the age (centered) covariate, and $\eta_{jm}\boldsymbol{=}0.02$ across all sites. Power curves for all methods from the main simulation are shown in Figure S7. Results do not differ substantially from those in the main paper, empirically verifying mixWAS’s robustness to missingness under MAR.

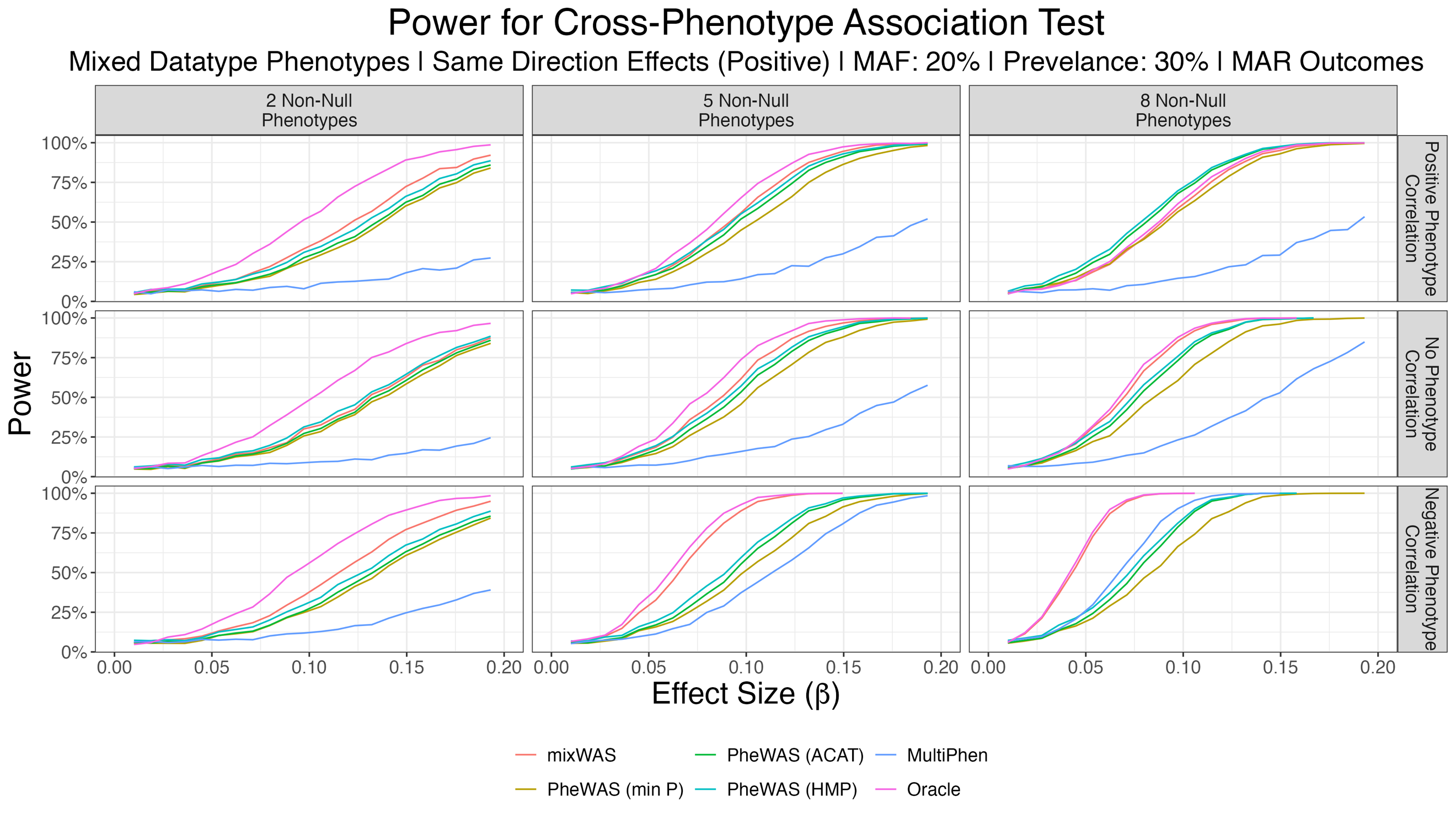

**Figure S7:** Empirical power curves comparing various cross-phenotype association tests for simulated phenotypes with outcomes missing at random. Missingness was driven my age, with older subjects more likely to display missingness across all phenotypes. This simulation setting is analogous to that presented in Figure 2A, with phenotypes MAR rather than MCAR.

**Heterogeneous Covariate Distributions**

Although previous simulation settings allowed covariate effects $\boldsymbol{\gamma}_{\boldsymbol{jm}}$ to differ across both sites and phenotypes, covariates were sampled from the same distribution across sites in all prior simulations. In this final simulation, we draw age (centered) and sex from heterogenous distributions across sites. In particular, we use the following hierarchical approach to sample simulated covariates.

Sex:

$$p_{\mathrm{sex}}\sim Uniform\left( 0.3, 0.7 \right)$$

$$Sex\sim Bernoulli(p_{\mathrm{sex}})$$

$$Age (centered):$$

$$p_{\mathrm{dist}}\sim Uniform\left( 0, 1 \right)$$

$$Z_{dist}\sim Bernoulli\left( p_{dist} \right)$$

$$\sigma_{age}\sim Uniform\left( 12, 18 \right)$$

$$Age (Centered) \sim\left\{ \begin{aligned} N(0, \sigma_{age)}^{2}, &Z_{dist}=1 \\ Gamma\left( 5, \frac{\sigma_{age}}{\sqrt{5}} \right)-\sqrt{5}\sigma_{age}, &Z_{dist}=0 \end{aligned} \right.$$

Using this sampling approach, we repeat the main simulation setting whose results are in Figure 2A. Results for heterogenous covariate distributions are shown in Figure S8. Empirical power curves do not differ significantly from the main simulation result, confirming that mixWAS is robust to heterogeneity across sites.

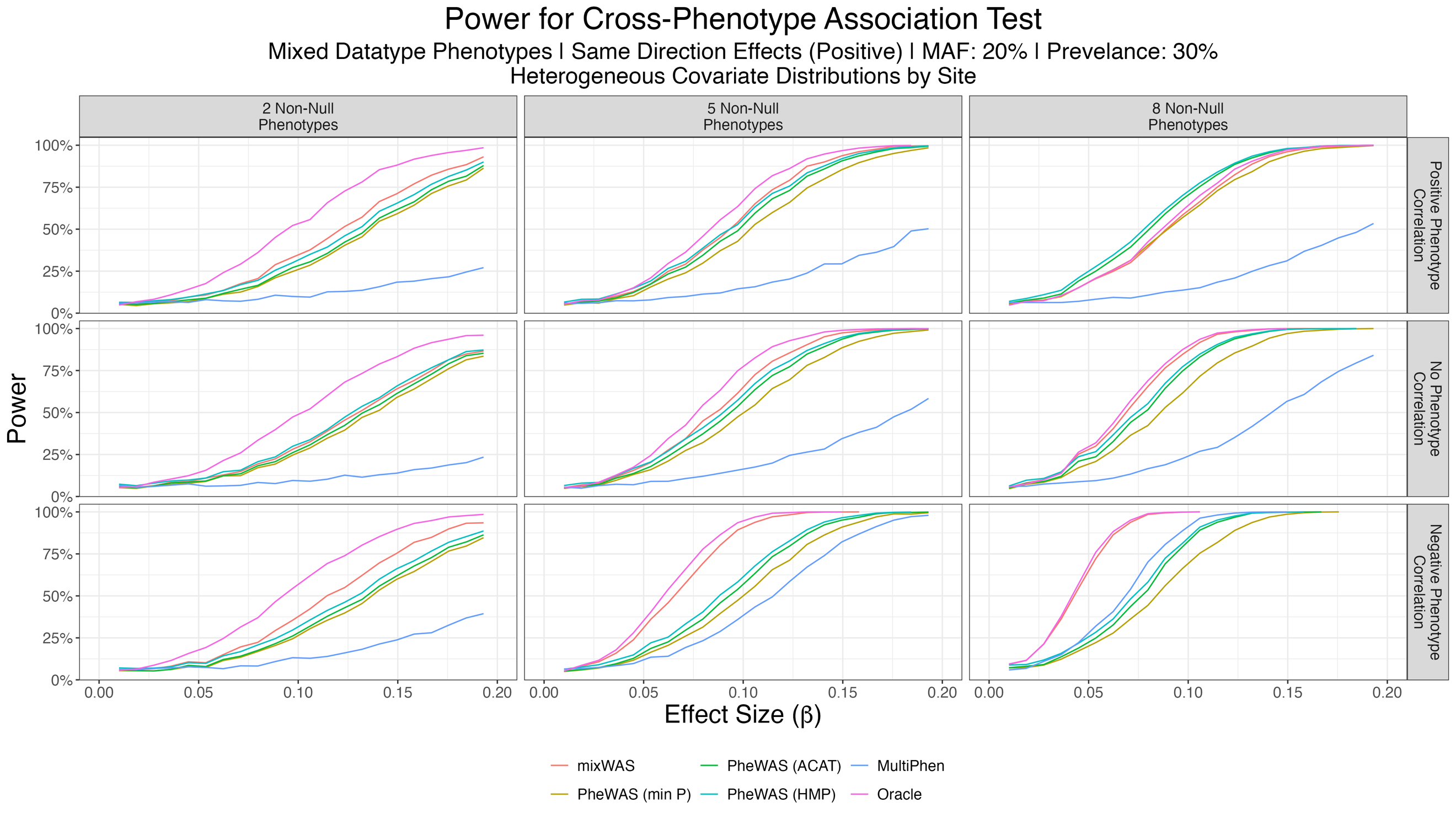

**Figure S8:** Empirical power curves comparing various cross-phenotype association tests for simulated phenotypes with heterogenous covariate distributions across sites.This simulation setting is analogous to that presented in Figure 2A, with the exception of how age (centered) and sex are sampled.

**mixWAS Algorithm**

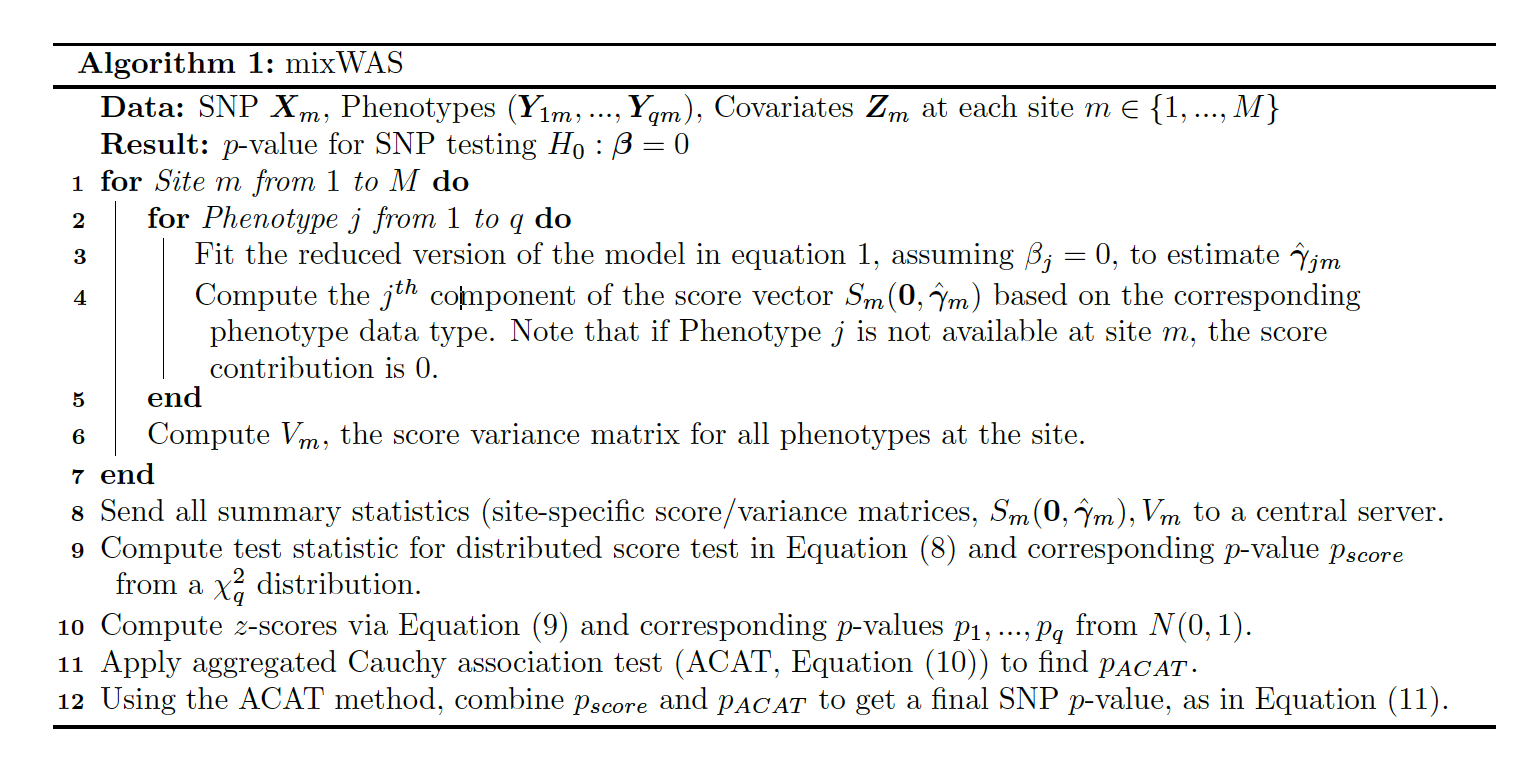

**Algorithm S1:** Outline of mixWAS algorithm
